# Population-scale multiome immune cell atlas reveals complex disease drivers

**DOI:** 10.1101/2025.11.25.25340489

**Authors:** Masahiro Kanai, Toni M Delorey, Jarno Honkanen, Rodosthenis S Rodosthenous, Julianna Juvila, Shane Murphy, Isabella Teixeira-Soldano, Hee Seung Hwang, Juha Karjalainen, Jussi Halonen, Georgia Panagiotaropoulou, Yuanxiang Zhang, Cristin McCabe, Eric Chen, Kosaku Nanki, Toshimi Yoshida, Kai Liu, Marla Glean, Nitya Mehrotra, Emily P Finan, Daniel Chafamo, Yixiao Zhu, Mikko Arvas, Sanni Ruotsalainen, Zhili Zheng, FinnGen, Mary P Reeve, Mitja Kurki, Caroline BM Porter, Orr Ashenberg, Wei Zhou, Kimmo Pitkänen, Jukka Partanen, Aarno Palotie, Daniel B Graham, Mark J Daly, Ramnik J Xavier

## Abstract

Most genetic variants associated with complex diseases lie in non-coding regions, yet mechanistic insights have been limited by the lack of an empirical framework for characterizing the molecular consequences of regulatory variation. Single-cell profiling of molecular quantitative trait loci (QTL) can connect variants to gene regulation, but prior studies lacked the sample size to detect variants at disease-relevant genes and the simultaneous measurements across regulatory layers needed to trace complete mechanisms from chromatin state to gene expression. Here we show that population-scale simultaneous profiling of chromatin accessibility and gene expression across immune cell types reveals multi-layered regulatory pathways connecting genetic variants to disease. We generated paired single-nucleus ATAC-seq and RNA-seq profiles from 10 million peripheral blood mononuclear cells across 1,108 Finnish individuals, identifying 51,083 *cis*-eQTLs for 20,829 genes, 338,100 *cis*-caQTLs for 210,584 peaks, 119,094 fine-mapped variants, and 496,488 enhancer–gene links. Systematic classification of regulatory mechanisms revealed that variants with complete chromatin-to-expression cascades show twice the disease colocalization of chromatin-only effects, establishing a hierarchy where mechanistic cascade predicts disease relevance. Analysis of evolutionarily constrained genes revealed multi-layered regulatory buffering where chromatin accessibility changes occur with normal effect sizes, but transmission to gene expression is attenuated through systematically weaker enhancer–gene links, reconciling why disease variants preferentially target these genes despite apparent eQTL depletion. We incorporated base editing to experimentally validate causal variants and mechanisms at Finnish-enriched disease loci such as *TNRC18*. This resource provides testable mechanistic hypotheses for over half of immune disease associations.

## Introduction

Genome-wide association studies (GWAS) have identified thousands of genetic loci associated with complex diseases^1,2^. However, most causal variants underlying these disease-associated loci cannot be fine-mapped to single variants^3^ and are located within non-coding regions^4,5^, limiting functional interpretation and experimental modeling. Molecular quantitative trait locus (molQTL) mapping, such as expression QTL (eQTL)^6–8^ or chromatin accessibility QTL (caQTL)^9,10^, has helped link variants to gene regulation. However, bulk approaches inherently obscure cellular heterogeneity by aggregating regulatory signals across heterogeneous cell populations, limiting power to detect cell type-or state-specific effects and leaving many disease-relevant regulatory mechanisms unexplained^11–13^.

Single-cell sequencing technologies now enable the study of genetic regulation at an unprecedented resolution^11–13^. Population-scale single-cell eQTL data has shown promise in elucidating molecular mechanisms across biological systems such as blood^14–18^, brain^19^, and skeletal tissue^20^. However, recent studies have revealed systematic differences between the genetic architecture of gene expression and complex traits, with expression-only approaches incompletely characterizing regulatory mechanisms and failing to effectively pinpoint causal genes^21–23^. Furthermore, only 11% of disease heritability is estimated to be mediated by *cis*-genetic components of gene expression^24^. Chromatin accessibility may better bridge the gap between genetic variation and gene regulation^21,25^, yet current single-cell studies are constrained by limited sample sizes and lack matching gene expression data to connect chromatin accessibility peaks to nearby genes^26,27^. Simultaneous profiling of gene expression and chromatin accessibility within the same nucleus^28,29^ enables more robust cell type identification, direct correlation between regulatory enhancer activity and gene expression in specific cell types, and linkage of variants to target genes through enhancer–gene associations^30–33^.

Here, we leverage single-nucleus paired multiome profiling to map regulatory variants across millions of immune cells in 1,108 peripheral blood mononuclear cell (PBMC) samples from participants in the FinnGen project^34^. This population-scale dataset provides the first opportunity to assess precision gains from multiome profiling when mapping thousands of common GWAS associations to effector genes. The Finnish population offers a distinct opportunity to study disease mechanisms through comprehensive population-wide longitudinal healthcare data and a strong population bottleneck ∼120 generations ago that stochastically enriched the frequency of certain disease variants^34^. The Finnish-enriched variants, despite their outsized impact on disease biology, are poorly understood because they are absent from existing multiomic datasets and must be characterized using Finnish samples. Importantly, the functional effects and regulatory mechanisms we characterize represent fundamental biology shared across human populations. Our framework thus elucidates mechanisms of Finnish-enriched variants while establishing generalizable principles for how common variants shape immune regulation, providing insights applicable to the genetic architecture of complex diseases across diverse populations.

By simultaneously measuring chromatin accessibility and gene expression at scale, we systematically map molQTLs and enhancer–gene links across immune cell types and develop a systematic framework for classifying regulatory mechanisms that reveals a clear hierarchy where mechanistic chromatin-to-expression cascades predict disease relevance. We show that caQTL variants capture substantially more disease-relevant regulation than eQTL variants alone and uncover multi-layered regulatory buffering at evolutionarily constrained genes that reconciles the long-standing paradox of why disease variants preferentially target these genes despite apparent eQTL depletion^8,21,22,35^. Colocalization with FinnGen GWAS traits demonstrates that the majority of immune-mediated disease associations can be attributed to regulatory mechanisms, with caQTLs showing higher colocalization rates than eQTLs. By integrating population-scale single-cell molQTL mapping with base-editing validation, we establish a variant-to-function framework connecting causal variants to biological functions, experimentally validating regulatory mechanisms of Finnish-enriched disease variants and demonstrating how human genetics and functional perturbation can be systematically combined to elucidate disease biology.

## Results

### Population-scale, single-nucleus gene expression and chromatin accessibility atlases

Through the Finnish Red Cross Blood Service^36^, we recruited 1,171 healthy Finnish donors enrolled in FinnGen (**Fig. 1a**). From each donor, we generated paired single-nucleus RNA-sequencing (snRNA-Seq) and single nucleus assay for transposase-accessible chromatin (snATAC-Seq) data from peripheral blood mononuclear blood cells (PBMCs; **Methods**). The first 339 donors were profiled in individual microfluidic Multiome assay channels (10x Genomics). To facilitate cohort scale-up, the remaining 832 samples were multiplexed in batches of 16 across eight channels, reducing both time and resource expenditure while maintaining high-quality data (**Extended Data Fig. 1a, Supplementary Table 1, Methods**).

**Fig. 1:**
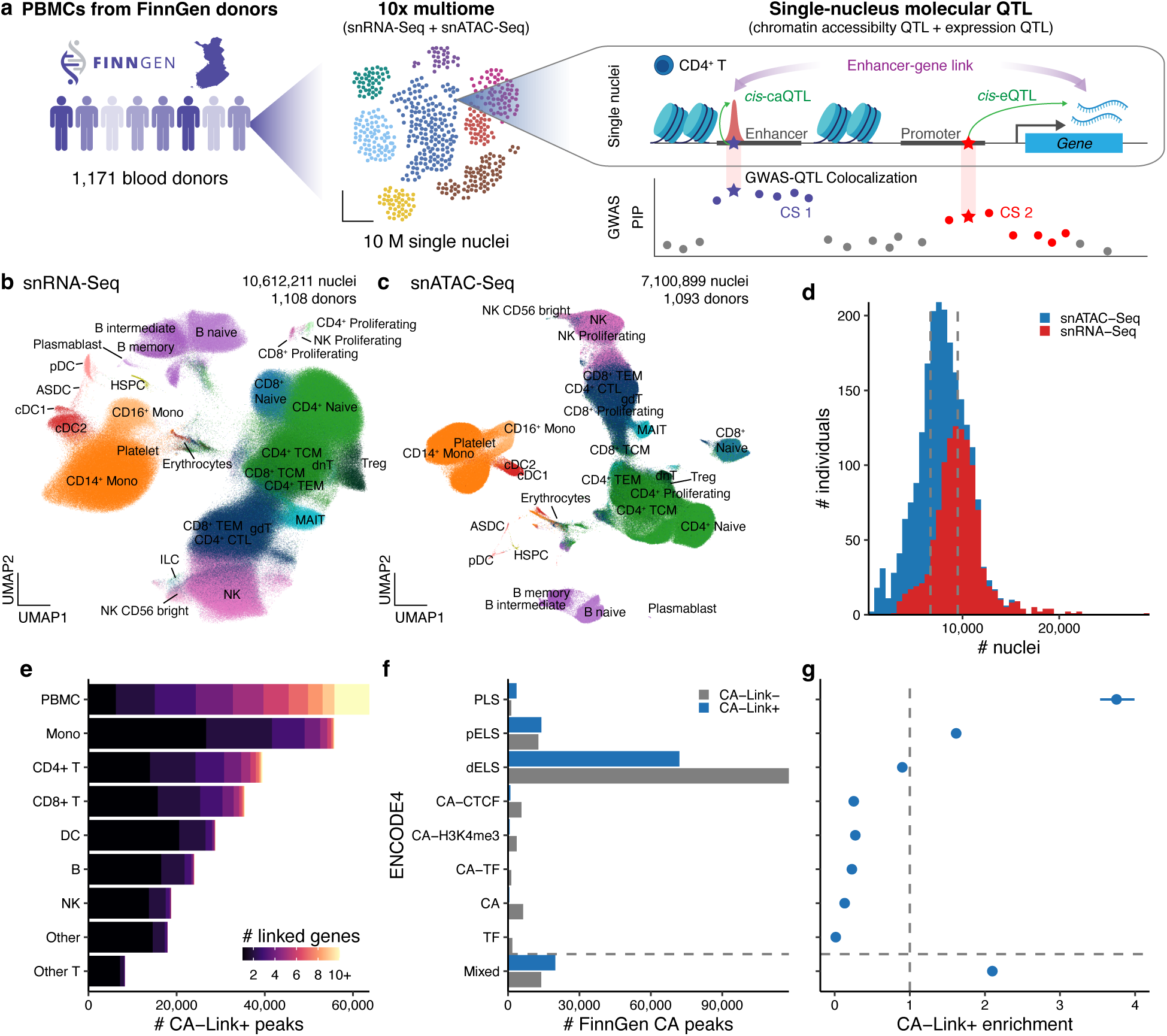
Single-nucleus RNA-seq and ATAC-seq atlases of 1,108 FinnGen donors. a. Schematic overview of the study. PBMC, peripheral blood mononuclear cell. QTL, quantitative trait locus. caQTL, chromatin accessibility QTL eQTL, expression QTL. GWAS, genome-wide association study. PIP, posterior inclusion probability. CS, credible set. b,c. UMAP representations of snRNA-Seq (n = 10,612,211 nuclei) and snATAC-Seq (n = 7,100.899 nuclei) atlases. Color represents L2 cell types (see Methods). d. The distribution of the number of nuclei per individual for snRNA-Seq and snATAC-Seq. Vertical lines represent the median of the number of nuclei per individual. e. The number of chromatin accessibility (CA) peaks with significant enhancer–gene links (CA–Link+; see Methods) across L1 cell types. Color represents the number of linked genes for each CA peak. Mono, monocyte. DC, dendritic cells. NK, natural killer cell. f. The number of FinnGen CA peaks for each ENCODE4 cCRE classifications, split by enhancer–gene link status (CA–Link– and CA–Link+). The mixed category represents the CA peak that overlaps with multiple cCRE classifications. g. The enrichment of CA–Link+ for each ENCODE4 cCRE classification. Enrichment is defined as a risk ratio of having a significant link (CA–Link+) and being in an annotation (see Methods). Error bars represent 95% confidence intervals.

We developed a computational quality control (QC) pipeline (**Extended Data Fig. 1b, Methods**) and generated a snRNA-seq atlas containing 10,612,211 high-quality nuclei (*n* = 1,108 donors) capturing 29,602 genes, with a median of 9,506 nuclei/donor, 1,056 genes/nucleus, and 1,828 unique molecular identifier (UMI)/nucleus (**Fig. 1b,d**, **Extended Data Fig. 2a,b**). Nuclei were annotated using label transfer from a single-cell RNA-seq PBMC atlas^37^ (Level 1 [L1]: nine broad cell types; Level 2 [L2]: 31 finer-grained cell types). The most frequent cell types recovered were CD14^+^ monocytes, CD4^+^ subsets (naive, T central memory), CD8^+^ subsets (naive, T effector memory), naive B cells, and natural killer (NK) cells (**Extended Data Fig. 2c, Supplementary Table 2**).

We next generated the snATAC-seq atlas containing 7,100,899 high-quality nuclei (*n* = 1,093), each with paired high-quality snRNA-Seq data, with a median of 6,692 nuclei/donor, 4,745 fragments/nucleus, and a transcription start site (TSS) enrichment score of 6.47 per nucleus (**Fig. 1c,d, Extended Data Fig. 2c–e**). We called L2 cell type-specific peaks using pseudobulk fragments with MACS2 (ref. ^38^) and defined 297,024 consensus peaks across cell types (**Extended Data Fig. 2f**, **Supplementary Table 3**, **Methods**). Overall, these atlases encompassed 1,108 QC-passing donors, 29,602 genes, and 297,024 consensus peaks for downstream analyses.

### Regulatory landscape of enhancer–gene links in *cis*

Correlations between chromatin accessibility and gene expression provide critical insights into enhancer–gene regulatory links^30–32^, pinpointing candidate regulatory peaks for individual genes. Leveraging our large-scale paired multiome measurements, we systematically mapped 496,488 enhancer–gene links with significant peak–gene associations in *cis* (i.e., < 1 Mb between TSS and peak) using a hurdle model to handle both zero-inflation and overdispersion in single-cell gene expression^32^ (**Fig. 1e**, **Supplementary Table 4**, **Methods**). These regulatory links encompass 112,032 consensus peaks (37.6%) that regulate 12,445 genes in *cis* across cell types (median 3 genes per peak, median 33 peaks per gene; **Extended Data Fig. 2g,h**), in addition to the remaining 184,992 consensus peaks that showed no conclusive links to nearby genes. Notably, we identified genes with extensive regulatory links at 19p13.11, including *IFI30*, *JUND*, *MAST3*, and *IL12RB1* (206, 187, 137, and 91 linked peaks, respectively; **Extended Data Fig. 3a**). This region contains zinc finger clusters that evolve rapidly through copy number variation to generate transcriptional repressors against endogenous retroviruses^39^. Similarly, TNF receptor superfamily (TNFRSF) members demonstrated remarkable regulatory complexity, particularly at the 8p21.3 cluster containing TRAIL receptors (*TNFRSF10A–D*), collectively regulated by 114 peaks (**Extended Data Fig. 3b**), consistent with their coordinated transcriptional regulation during apoptosis and immune homeostasis. These regulatory hotspots exemplify how clustered gene families with shared functions exhibit exceptionally complex regulatory architectures.

To comprehensively characterize our chromatin accessibility peaks, we integrated ENCODE4 candidate *cis*-regulatory elements (cCREs) as an orthogonal reference^40^. The majority of peaks were annotated as distal enhancer-like signatures (dELS, 63.7%), while proximal enhancer-like signatures (pELS, 9.0%) and promoter-like signatures (PLS, 1.7%) showed the strongest enrichment for enhancer–gene links (risk ratio [RR] = 3.84 and 1.60, respectively; **Fig. 1g**, **Supplementary Table 5, Methods**), demonstrating that promoter-proximal cCREs are disproportionately enriched for our peak–gene links in *cis*. This comprehensive map of *cis*-regulatory links enables direct tracing of regulatory cascades from chromatin accessibility to gene expression, serving as the foundational resource for subsequent analyses.

### Genetic architecture of molecular traits in PBMCs

To investigate the genetic architecture of gene expression and chromatin accessibility in PBMCs, we tested associations of 17,167,198 imputed genetic variants with 27,294 genes and 297,024 chromatin accessibility peaks that are expressed or accessible in at least one of nine L1 and 28 L2 cell types (**Fig. 1b**, **Methods**).

We first tested associations between gene expression and genetic variants located in *cis* (*i.e.*, < 1 Mb within TSS; **Methods**) using SAIGE-QTL^16^, which enables single-nucleus eQTL mapping based on a Poisson mixed model. We identified 20,829 unique genes (76% of tested genes; FDR < 0.05; *cis*-eGenes) whose expression is modified by *cis*-genetic variants in at least one cell type (**Fig. 2a, Extended Data Fig. 4a, Supplementary Table 6**). Statistical fine-mapping of *cis*-eQTL associations identified 86,536 and 89,350 independent 95% credible set (CS)–gene–cell type trios for L1 and L2 cell types, respectively, with 16,477 unique fine-mapped *cis*-eQTL variants with posterior inclusion probability (PIP) > 0.5 (**Fig. 2b, Extended Data Fig. 4b, Supplementary Table 7**). To assess sharing of *cis*-eQTLs across cell types, we colocalized CSs for the same gene across cell types, identifying 51,083 merged CS–gene pairs from 175,886 CS–gene–cell type trios, with CSs showing higher colocalization within related immune cell types (**Extended Data Fig. 4c**, **Supplementary Table 8**, **Methods**). To benchmark our results, we separately tested pseudobulk *cis*-eQTL associations using tensorQTL^41^, and observed that SAIGE-QTL had an average 18% increase in the number of eGenes (**Extended Data Fig. 5a, Supplementary Table 9**).

**Fig. 2:**
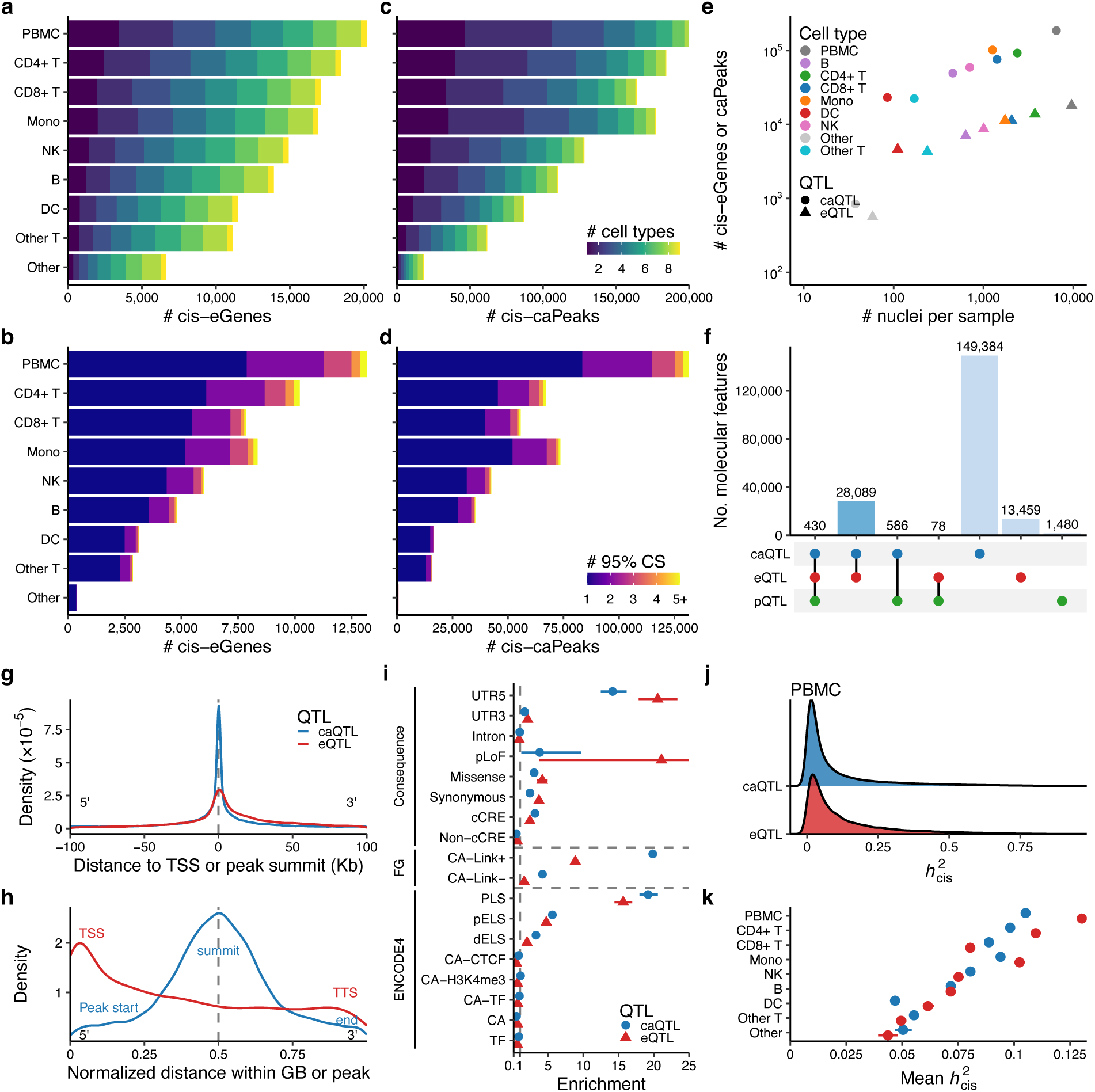
Genetic architecture of molecular QTLs in FinnGen. a,c. The number of significant *cis*-eGenes (**a**) and *cis*-caPeaks (**c**) for each L1 cell type (FDR < 0.05; see **Methods**). Color represents the number of cell types that each eGene or caPeak is significantly detected for. **b,d**. The number of *cis*-eGenes (**b**) and *cis*-caPeaks (**d**) with fine-mapped 95% CSs. Color represents the number of 95% CSs for each *cis*-eGene or *cis*-caPeak. **e**. The relationship between the number of *cis*-eGenes or caPeaks and the number of nuclei per sample. Color represents different L1 cell types and shape represents either caQTL or eQTL. **f**. The number of molecular features for each cluster after clustering 95% CSs across caQTL, eQTL, and pQTL (see **Methods**). **g**. The distribution of fine-mapped eQTL or caQTL variants around transcription start site (TSS) or peak summit, respectively. For each 95% CS in PBMC, the variant with the highest PIP was extracted (see **Methods**). Negative distances are towards 5’ end and positive distances are towards 3’ end. For eQTL, distances are aligned to each gene’s strand. For caQTL, distances are computed on the positive strand. **h**. The distribution of fine-mapped eQTL or caQTL variants within gene body (GB) or peak. For eQTL, the normalized distances are computed between transcription start site (TSS) and transcription termination site (TTS). For caQTL, the normalized distances are computed between peak start and peak end, where peak summit is the center of the peak. For the sake of simplicity, we only retained the peak of 500 bp length for this analysis (see **Methods**). **i**. Functional enrichment of high-PIP (> 0.9) caQTL or eQTL variants. Enrichment is defined as a risk ratio of being fine-mapped and being in an annotation (see **Methods**). Error bars represent 95% confidence intervals. **j**. The distribution of estimated cis heritabilities for caQTL and eQTL in PBMC. **k**. The distribution of mean estimated *cis*-heritabilities for caQTL and eQTL across L1 cell types. The y-axis is ordered by the number of eGenes for each cell type using the same order as in (**a**). Error bars represent 95% confidence intervals.

To identify chromatin accessibility peaks that are modified by genetic variants in *cis*, we tested associations between chromatin accessibility peaks and *cis*-variants within 100 kb of peaks at pseudobulk level using tensorQTL^41^ to account for inherent sparsity of snATAC-seq fragments (**Methods**). We identified 210,584 unique chromatin accessibility peaks (71% of tested peaks; FDR < 0.05; *cis*-caPeaks) whose accessibility is genetically regulated by *cis*-variants in at least one cell type (**Fig. 2c, Extended Data Fig. 4d, Supplementary Table 10**). Fine-mapping of *cis*-caQTL associations identified 636,566 and 506,183 independent 95% CS–peak–cell type trios for L1 and L2 cell types, respectively, with 109,163 unique fine-mapped *cis*-caQTL variants with PIP > 0.5 (**Fig. 2d, Extended Data Fig. 4e, Supplementary Table 11**). Colocalization of *cis*-caQTL CSs for the same peak across cell types identified 338,100 merged CS–peak pairs from 1,142,749 CS–peak–cell type trios (**Extended Data Fig. 4f, Supplementary Table 12**).

We observed that the number of eGenes and caPeaks scales with the number of nuclei per sample, with eQTLs approaching saturation faster compared to caQTLs (**Fig. 2e**; **Extended Data Fig. 4g,h, Supplementary Table 13**). This power relationship is partially explained by sparsity, as the number of tested genes or peaks also follows a similar scaling with the number of nuclei per sample (**Extended Data Fig. 4i**).

Colocalization across molQTLs identified extensive sharing of genetic signals across regulatory layers, with notable differences in their propagation patterns. We further incorporated plasma *cis*-protein QTLs (*cis*-pQTLs) from FinnGen (Olink Explore 3072, *n* = 1,732, **Methods**) to analyze genetic effects beyond transcriptional and chromatin regulation. To systematically identify CSs that affect multiple peaks, genes, and proteins, we performed colocalization among all *cis*-caQTLs, eQTLs, and pQTLs, identifying 193,506 merged CSs across molQTLs and cell types (**Fig. 2f, Supplementary Table 14**). Of these, 28,089 CSs (15%) showed colocalization between caQTLs and eQTLs, with a median of three caPeaks colocalizing with each eGene. Only 430 CSs (0.2%) exhibited colocalization across all three molecular layers (caQTLs, eQTLs, and pQTLs), partly because only 9% of the 27,294 tested genes had corresponding proteins measured by Olink in plasma, with 217 of these CSs regulating the same gene and its protein product. For example, we identified an intronic variant of *LILRB4*, rs776736:A>G, that increases *LILRB4* gene expression in monocytes (*β* = 0.16, *P* = 1.7 × 10^-25^, PIP = 1.0) and protein expression (*β* = 0.34, *P* = 6.5 × 10^-16^, PIP = 1.0) in plasma (**Extended Data Fig. 5b**). LILRB4 is an inhibitory immune checkpoint receptor expressed primarily on monocytes and dendritic cells that suppresses T cell activation, making it a potential therapeutic target for cancer immunotherapy and autoimmune diseases^42^. The majority of CSs colocalized only within each molQTL: 149,384 (77%), 13,459 (7%), and 1,480 (0.8%) CSs for caQTL, eQTL, and pQTL alone, respectively. These patterns of colocalization reveal that molQTLs operate through both coordinated effects across molecular layers and layer-specific mechanisms, highlighting the complex relationship between chromatin state, gene expression, and protein abundance.

Examination of genetic architecture revealed that both *cis*-eQTLs and *cis*-caQTLs were predominantly local, with median distance to TSS of 5,371 bp and to peak summits of 249 bp, respectively (**Fig. 2g, Extended Data Fig. 5c,d**). Within individual gene bodies or peaks, *cis*-eQTLs were localized most frequently at transcription start sites (TSS), whereas *cis*-caQTLs were localized at peak summits, reflecting their critical positions for regulating transcription and chromatin accessibility, respectively (**Fig. 2h**).

To functionally categorize *cis*-eQTL and *cis*-caQTL variants, we systematically evaluated the functional enrichment of high-PIP (> 0.9) fine-mapped molQTL variants across functional consequences (predicted loss-of-function [pLoF], missense, synonymous, 5’/3’ untranslated region [UTR]), ENCODE4 cCRE classifications, and our peak–gene link annotations (CA-Link+ and CA-Link–). Both eQTL and caQTL variants showed strongest enrichment in 5’ UTRs (RR = 21 and 14, respectively), followed by 3’ UTRs (RR = 2.1 and 1.7, respectively; **Fig. 2i, Supplementary Table 15**). While coding variants (missense and synonymous) showed moderate enrichment (RR = 3.7–4.2 and 2.4–3.0 for eQTL and caQTL, respectively), this was largely driven by variants overlapping cCREs: coding variants without cCRE overlap showed substantially reduced enrichment compared to those with cCRE overlap (RR = 1.1–2.7 without vs. 4.7–6.8 with cCRE overlap; **Extended Data Fig. 5e, Supplementary Table 16**).

Fine-mapped molQTL variants were dramatically enriched for chromatin accessibility peaks with enhancer–gene links (CA-Link+), particularly for caQTLs (RR = 20) compared to eQTLs (RR = 8.9), while peaks without such links (CA-Link–) showed modest enrichment (**Fig. 2i, Supplementary Table 15**). Among ENCODE4 cCRE classifications, PLS exhibited the highest enrichment (RR = 16 and 19 for eQTL and caQTL, respectively), followed by pELS (RR = 4.7 and 5.6, respectively) and dELS (RR = 2.0 and 3.3, respectively), while CA-only elements showed depletion (RR = 0.49–0.69 and 0.51–1.1, respectively). These functional enrichment patterns underscore the regulatory nature of most molQTL variants and highlight the utility of our enhancer–gene links for nominating functional variants.

The mean narrow-sense *cis*-heritabilities (*h*^2^_cis_) of gene expression and chromatin accessibility were estimated as 0.13 and 0.11 for PBMCs, with approximately 1.0% and 5.1% of each gene’s or peak’s variance explained by fine-mapped *cis*-eQTL or *cis*-caQTL variants on average, respectively (**Fig. 2j**, **Methods**). The mean *h*^2^_cis_ approximately scales with statistical power across cell types, recapitulating genetic architecture observed for complex traits (**Fig. 2k, Supplementary Table 17**).

Finally, we also tested associations between molecular traits and genetic variants located in *trans* (*trans*-eQTLs, > 5 Mb away or on different chromosomes; **Methods**) using tensorQTL^41^ to explore distal genetic regulation (**Extended Data Fig. 5f,g**). We identified 20,826 unique *trans*-eGenes and 215,949 unique *trans*-caPeaks in at least one cell type (*P* < 5.0 × 10^-8^; **Supplementary Table 18,19, Methods**), of which 85% and 76% are also *cis*-eGenes and *cis*-caPeaks, respectively, suggesting the majority of genes and peaks are both *cis*-and *trans*-regulated. Notably, among 86,536 *cis*-eQTL and 636,566 *cis*-caQTL 95% CS-gene-cell type trios, only 2,704 (3.1%) and 37,916 (6.0%) trios showed at least one significant *trans*-eQTL or *trans*-caQTL association with another gene or peak for the same cell type, respectively, suggesting distinct genetic regulation of *cis* and *trans* components.

### Cell type specificity of molecular QTLs

To characterize cell-type specificity and regulatory mechanisms of molQTLs, we developed a systematic framework, CASCADE (Comprehensive Analysis Suite for Cell-type specificity and Accessibility-Driven QTL Effects). Here, we first applied CASCADE to define hierarchical cell type specificity based on cell lineages and leverage multivariate adaptive shrinkage (mash)^43^ to identify likely shared but underpowered genes or peaks (**Methods**). We classified eGenes and caPeaks into five categories: 1) cross-lineage shared, with significant effects across myeloid and lymphoid lineages; 2) likely shared but underpowered, significant in one lineage (or only PBMC) but with low local false sign rate (LFSR) across lineages; 3) lineage-specific, restricted to either myeloid or lymphoid cell types; 4) T-cell-specific; and 5) single cell-type specific (**Fig. 3a**, **Methods**).

**Fig. 3:**
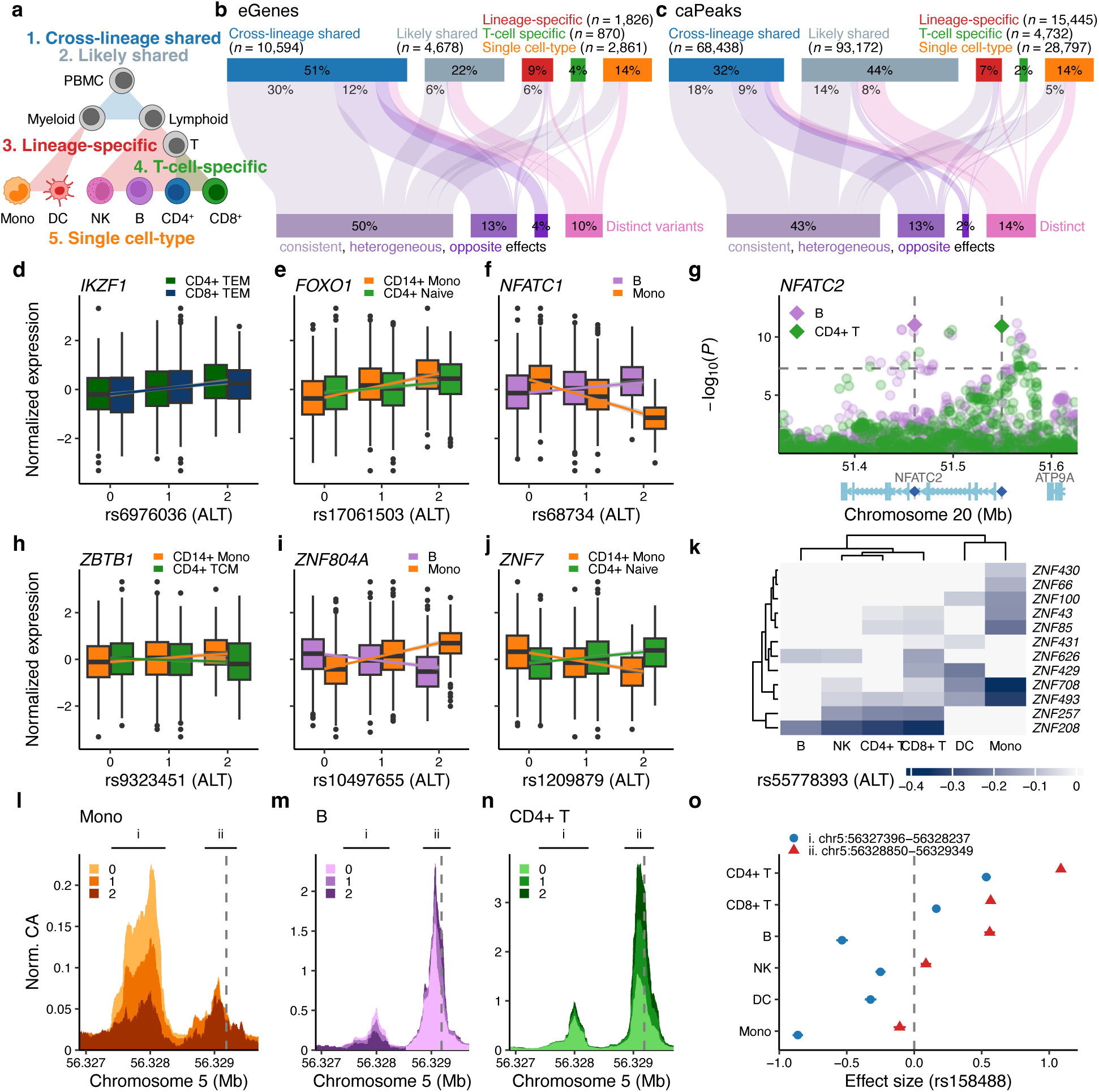
Cell type specificity of molecular QTLs. **a.** Schematic overview of cell type specificity definitions by CASCADE (see **Methods**). **b,c**. Cell-type specificity classifications of eGenes (**b**) and caPeaks (**c**). The top bar represents cell-type specificity, whereas the bottom bar represents whether shared eGenes or caPeaks across cell types arise from the same or distinct causal variants (see **Methods**). **d–f**. Examples of eGenes that are driven by the same variant with consistent (**d**, *IKZF1*), heterogeneous (**e**, *FOXO1*), and opposite (**f**, *NFATC1*) effects across cell types. Effect sizes are with respect to an alternative allele (ALT). **g**. An example of eGene, *NFATC2*, is driven by distinct causal variants. Horizontal dotted line represents the genome-wide significance threshold (*P* < 5.0 × 10^-8^). **h–j**, *ZNF* eGenes with opposite effects across cell types. **k**. Effect sizes of rs55778393 across cell types for *ZNF* cluster genes on chromosome 19. **l–n**, snATAC-seq coverage tracks, stratified by rs158488 genotypes. Sequence coverage (chromatin accessibility, CA) is normalized per one million fragments. Peak locations of peak i (chr5:56327396-56328237) and peak ii (chr5:56328850-56329349) are annotated at the top of each plot. Colors represent the number of alternative alleles of rs158488. The vertical dotted line represents the location of rs158488. **o**. Forest plot of caQTL effect sizes of rs158488 for peak i and ii across L1 cell types. Effect sizes are with respect to an alternative allele.

Among 20,829 eGenes, 73% showed shared regulation (51% cross-lineage and 23% likely shared but underpowered), while 27% exhibited cell type specific patterns (9% lineage-specific, 4% T-cell-specific, and 14% single cell-type; **Fig. 3b, Supplementary Table 20**). Chromatin accessibility showed comparable sharing: of 210,584 caPeaks, 77% showed sharing (33% cross-lineage and 44% likely shared), with 23% displaying cell type specific patterns (7% lineage-specific, 2% T-cell-specific, and 14% single cell-type; **Fig. 3c, Supplementary Table 21**). The large proportion of likely shared but underpowered caPeaks (44%) suggests that the genetic regulation of chromatin accessibility is broadly shared across immune cell types, with observed specificity solely based on significance often reflecting limited statistical power rather than true biological differences.

We next examined whether shared eGenes or caPeaks across cell types arise from the same or distinct causal variants, analyzing those with sharing across multiple cell types (excluding single cell-type) and with fine-mapped CSs (**Methods**). We were able to classify 67% of eGenes and 58% of caPeaks as driven by at least one of the same causal variants across cell types, whereas 10% of eGenes and 14% of caPeaks involved distinct causal variants, while the remainder were unclassified. Among eGenes and caPeaks with shared causal variants, we further characterized their effect consistency using Cochran’s *Q* heterogeneity test (*P* < 5.0 × 10^-8^). Specifically, 50% of eGenes and 43% of caPeaks showed consistent effects, while 13% and 13% showed heterogeneous effects (with the same direction), and 4% and 2% exhibited opposite effects despite sharing the same causal variants (**Fig. 3b,c, Supplementary Table 20,21**).

These variant effect patterns manifested in transcription factors (TFs) with clear immunological relevance. The lymphoid-specific *cis*-eQTL for *IKZF1* (rs6976036:T>C) showed consistent effects in CD4^+^ and CD8^+^ T cells (*β* = 0.04 and 0.04, *P* = 8.1 × 10^-34^ and 8.4 × 10^-21^, respectively; **Fig. 3d**). *IKZF1* encodes Ikaros, a master TF essential for lymphoid cell development that regulates B, T, and NK cell differentiation throughout hematopoiesis. The *cis*-eQTL for *FOXO1* (rs5803064:ACTT>A) exhibited heterogeneous cross-lineage effects, with stronger regulation in CD14^+^ monocytes than CD4^+^ naive T cells (*β* = 0.11 and 0.03, *P* = 2.2 × 10^-94^ and 5.1 × 10^-15^, Cochran’s *Q p*-value = 3.1 × 10^-365^; **Fig. 3e**). *FOXO1* is forkhead box TF with prominent roles in monocyte inflammatory responses, while in T cells it primarily maintains homeostasis through IL-7 receptor regulation. Strikingly, *NFATC1* showed opposite effects between monocytes and B cells for the same variant (rs68734:G>C; *β* = –0.18 and 0.07, *P* = 3.8 × 10^-89^ and 8.0 × 10^-17^, respectively; **Fig. 3f**), while *NFATC2* was regulated by distinct variants in different cell types (rs4809845:T>C in B cells and rs6021282:T>A in CD4^+^ T cells; **Fig. 3g**), both of which encode NFAT transcription factors that regulate immune cell activation and differentiation.

Zinc finger (ZNF) genes represent a particularly intriguing class of immune regulators, as many remain functionally uncharacterized despite growing evidence linking them to immune disorders and broader biological processes. Among 375 ZNF genes analyzed (including *ZNF*, *ZBTB*, *ZSCAN*, *ZKSCAN*, and *ZFP* families; **Methods**), 345 (92%) were identified as eGenes. Of these 271 showed shared regulation (221 cross-lineage and 51 likely shared but underpowered), while 74 exhibited cell-type specific patterns (38 lineage-specific, nine T-cell-specific, and 24 single cell-type). For example, a 5’ UTR variant of *ZBTB1*, rs9323451:G>A, decreased *ZBTB1* expression in CD4^+^ central memory T cells while increasing it in CD14^+^ monocytes (*β* = 0.05 and –0.02, *P* = 3.8 × 10^-7^ and 8.2 × 10^-6^, respectively; **Fig. 3h**), which is notable given that *ZBTB1* knockout mice exhibit T cell deficiency^44^. Similarly, the *cis*-eQTL for *ZNF804A* (rs10497655:T>C) decreased *ZNF804A* expression in B cells but increased expression in monocytes (*β* = –0.18 and 0.13, *P* = 1.0 × 10^-17^ and 4.6 × 10^-87^, respectively; **Fig. 3i**), while the *cis*-eQTL for *ZNF7* (rs1209879:C>T) decreased *ZNF7* expression in CD14^+^ monocytes and increased in CD4^+^ naive T cells (*β* = –0.16 and 0.06, *P* = 2.0 × 10^-45^ and 3.3 × 10^-11^, respectively; **Fig. 3j**), demonstrating opposing effects between lymphoid and myeloid lineages. Notably, we identified variants that regulate multiple clustered *ZNF* genes, particularly on chromosome 19 where single variants, such as rs55778393:C>A, controlled expression of up to 12 co-localized *ZNF* genes at 19p12 (**Fig. 3k**). This coordinated genetic regulation of ZNF clusters complements the extensive peak-gene links at these loci (**Extended Data Fig. 3c**), revealing how architectural complexity and genetic co-regulation together shape zinc finger repertoire diversity^39^.

Similarly, chromatin accessibility revealed how genetic variants differentially affect regulatory elements across cell types. At the *IL6ST-ANKRD55-MAP3K1* locus, the same variant rs158488:G>A regulated two adjacent peaks with contrasting patterns: peak i (chr5:56,327,396–56,328,237) showed negative regulation in monocytes and B cells but positive in T cells, while peak ii (chr5:56,328,850–56,329,349) was positively regulated only in B and T cells (**Fig. 3l–o**). This demonstrates how identical genetic variants can exert opposite regulatory effects depending on cellular context, providing mechanistic insights into cell type-specific gene regulation.

### Genetic regulation of chromatin accessibility and gene expression cascades

To systematically characterize how genetic variants regulate gene expression through chromatin accessibility, we next applied CASCADE to 119,094 fine-mapped molQTL variants (PIP > 0.5), examining their regulatory mechanisms across PBMC cell types. We classified each variant into one of 25 mutually exclusive patterns based on four key features: i) chromatin accessibility peak overlap; ii) caQTL status; iii) peak–gene links; and iv) eQTL status (**Fig. 4a**, **Extended Data Fig. 6a**, **Methods**).

**Fig. 4:**
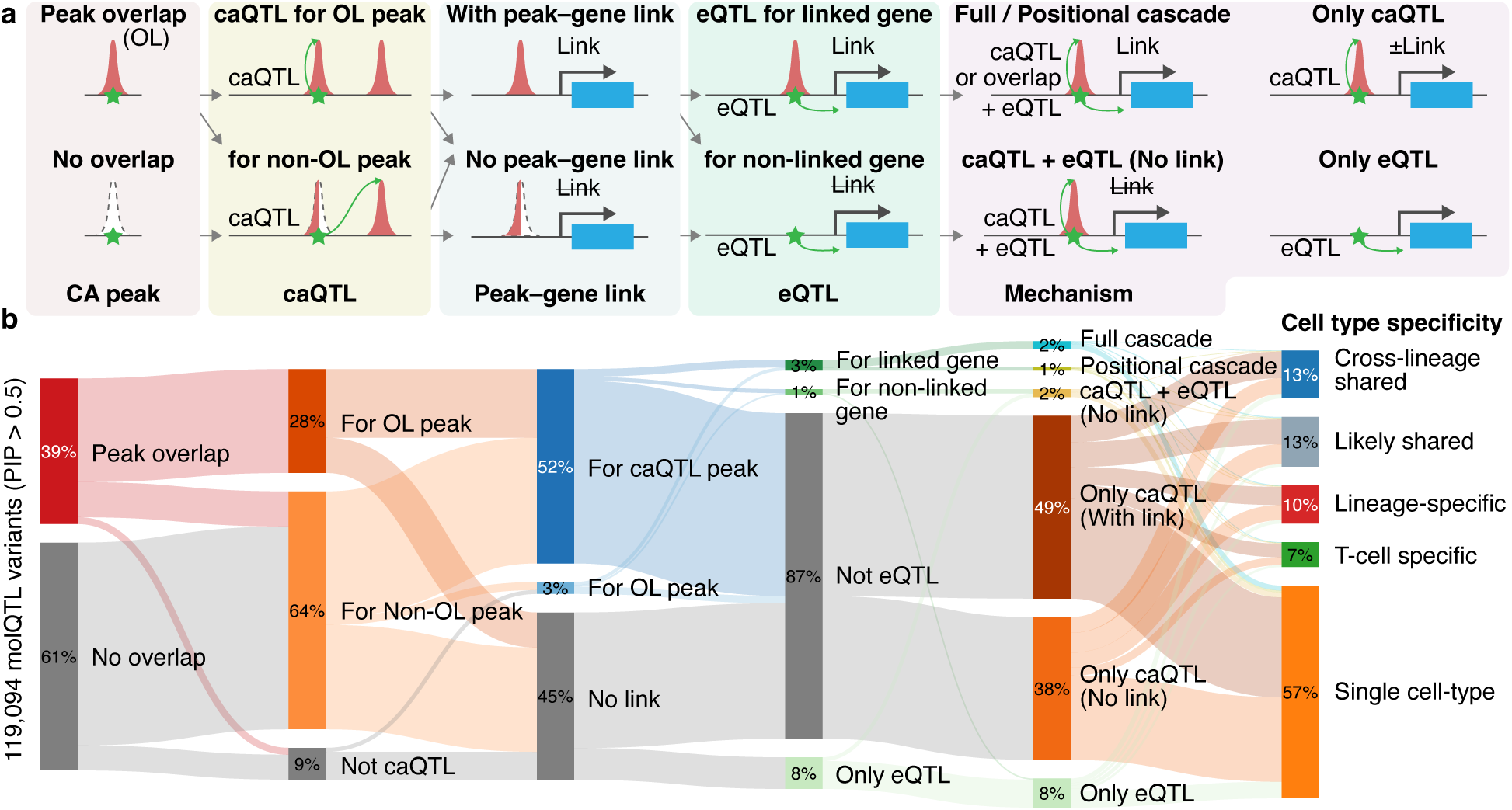
Comprehensive classification of QTL mechanisms for fine-mapped molQTL variants. **a.** Schematic overview of QTL mechanism classification workflow by CASCADE (see **Methods**). Each variant is classified into one of 25 mutually exclusive patterns based on i) chromatin accessibility (CA) peak overlap, ii) caQTL, iii) peak–gene link, and iv) eQTL, then grouped into six mechanism categories, i.e., full cascade, positional cascade, caQTL + eQTL (no link), only caQTL (with and without link), and only eQTL (see **Methods**). **b**. Sankey diagram of the QTL mechanism classifications of 119,094 fine-mapped molQTL variants (PIP > 0.5). For each layer, nodes represent proportions of the fine-mapped molQTL variants. Edges between nodes in each layer represent the fraction of variants shared between nodes.

Among all analyzed variants, 39% overlapped chromatin accessibility peaks, of which 71% (28% of all variants) showed caQTL effects for those overlapping peaks (**Fig. 4b, Supplementary Table 22**). Conversely, among the 61% of variants not overlapping peaks, 89% (54% of all variants) affected non-overlapping peaks as caQTLs. An additional 10% of all variants overlapped peaks but affected different, non-overlapping peaks, demonstrating complex accessibility regulation. Strikingly, 55% of molQTL variants have enhancer–gene links (18% via overlapping peak caQTL, 34% via non-overlapping peak caQTL, and 3% via peak overlap alone), highlighting functional connectivity between chromatin and gene regulation.

The 25 patterns collapsed into six mechanistic categories (**Fig. 4a**). Full cascades (complete regulatory pathways from caQTL through peak–gene links to eQTL) comprised 2% of fine-mapped molQTL variants (*n* = 2,429). Positional cascades, where variants overlap peaks with peak–gene links and show eQTL effects for the linked gene but no detectable caQTL, represented 1% (*n* = 995), likely affecting TF binding or other regulatory mechanisms. Another 2% (*n* = 2,535) showed both caQTL and eQTL effects but lacked connecting peak–gene links, suggesting either incomplete regulatory mapping or genuine pleiotropic effects. The vast majority exhibited partial effects: 87% demonstrated only caQTL (49% with links and 38% without), while 8% showed only eQTL effects (**Fig. 4b, Supplementary Table 22**). These fractions change substantially when analyzing only fine-mapped eQTL variants (*n* = 16,477), where 23% demonstrate full or positional cascades, 17% showed both caQTL and eQTL effects without links, and 61% remain as only eQTL variants, reflecting the excess of only caQTL variants (*n* = 103,851) in our fine-mapped molQTLs (**Extended Data Fig. 6b**).

A canonical example of a full cascade variant is rs72928038:G>A, a known risk factor for autoimmune diseases while protecting against skin cancer^45,46^, which regulates *BACH2* expression through chromatin accessibility. Located within an enhancer region^40^, this variant significantly decreases chromatin accessibility of its overlapping peak in CD4^+^ T cells (chr6:90266796-90267690; *β* = –1.3, *P* = 2.4 × 10^-268^, PIP = 1.0) that is strongly linked to *BACH2* expression (*β* = 0.20, *P* = 1.3 × 10^-1213^), thereby reducing *BACH2* expression (*β* = –0.09, *P* = 9.2 × 10^-48^, PIP = 0.61) in a full regulatory cascade (**Extended Data Fig. 6c**). Of note, another variant in LD, rs6908626:G>T (*r*^2^ = 0.99) in the *BACH2* promoter, also shows apparent but weaker chromatin accessibility effects on its overlapping peak in CD4^+^ T cells (chr6:90295895-90296663; *β* = –0.21, *P* = 4.2 × 10^-10^, PIP = 0.53) that is linked to *BACH2* expression (*β* = 0.05, *P* = 2.0 × 10^-130^; **Extended Data Fig. 6d**). However, the enhancer and promoter peaks demonstrate significant co-accessibility even in individuals lacking both variants (Pearson’s *r* = 0.20, *P* = 2.0 × 10^-9^; **Methods**), suggesting that rs6908626’s apparent effect likely reflects LD with rs72928038, whose impact propagates from enhancer to promoter through their physical interaction as demonstrated by promoter capture Hi-C^47,48^. This example, supported by previous MPRA validation^49^, illustrates how regulatory cascades operate through coordinated chromatin domains and underscores the need to fine-map co-accessible peaks jointly, as single variant effects can propagate through co-accessible peaks.

Variant-level cell type specificity analysis revealed striking heterogeneity across mechanisms (**Fig. 4b, Supplementary Table 22**). Overall, 26% of variants showed shared effects across cell types (13% cross-lineage and 13% likely shared), whereas 74% displayed cell type specific patterns (10% lineage-specific, 7% T-cell-specific, and 57% single cell-type). Full cascade variants were slightly more cell type-specific (**Extended Data Fig. 6e**): 20% were shared (7% cross-lineage and 13% likely shared) and 80% were cell type-specific (10% lineage-specific, 8% T-cell-specific, 63% single cell-type), suggesting that individual regulatory variants tend to operate through cell-type specific mechanisms even though eGenes and caPeaks themselves are largely shared across cell types.

### Leveraging molecular QTLs for GWAS colocalization

Having established a compendium of molQTLs in PBMCs, we performed colocalization analysis between molQTLs and GWAS signals across a wide range of complex diseases and traits to better prioritize putative causal genes and molecular mechanisms. Among 31,983 95% CS–trait pairs from 439 complex trait GWAS conducted in FinnGen (**Supplementary Table 23**), we identified 17,317 pairs (54%) where the same causal variant likely explains both the GWAS and the molQTL association (posterior probability for colocalization, PP.H4 > 0.5; **Methods**). Consistent with their upstream regulatory role, caQTLs exhibited higher colocalization rates (51.7%) than eQTLs (27.4%; **Fig. 5a**). From the perspective of molecular traits, this translates to 14% of 208,659 peaks and 24% of 20,598 genes tested for colocalization had at least one GWAS colocalization, indicating that a substantial fraction of genetically regulated molecular traits are directly relevant to diseases.

**Fig. 5:**
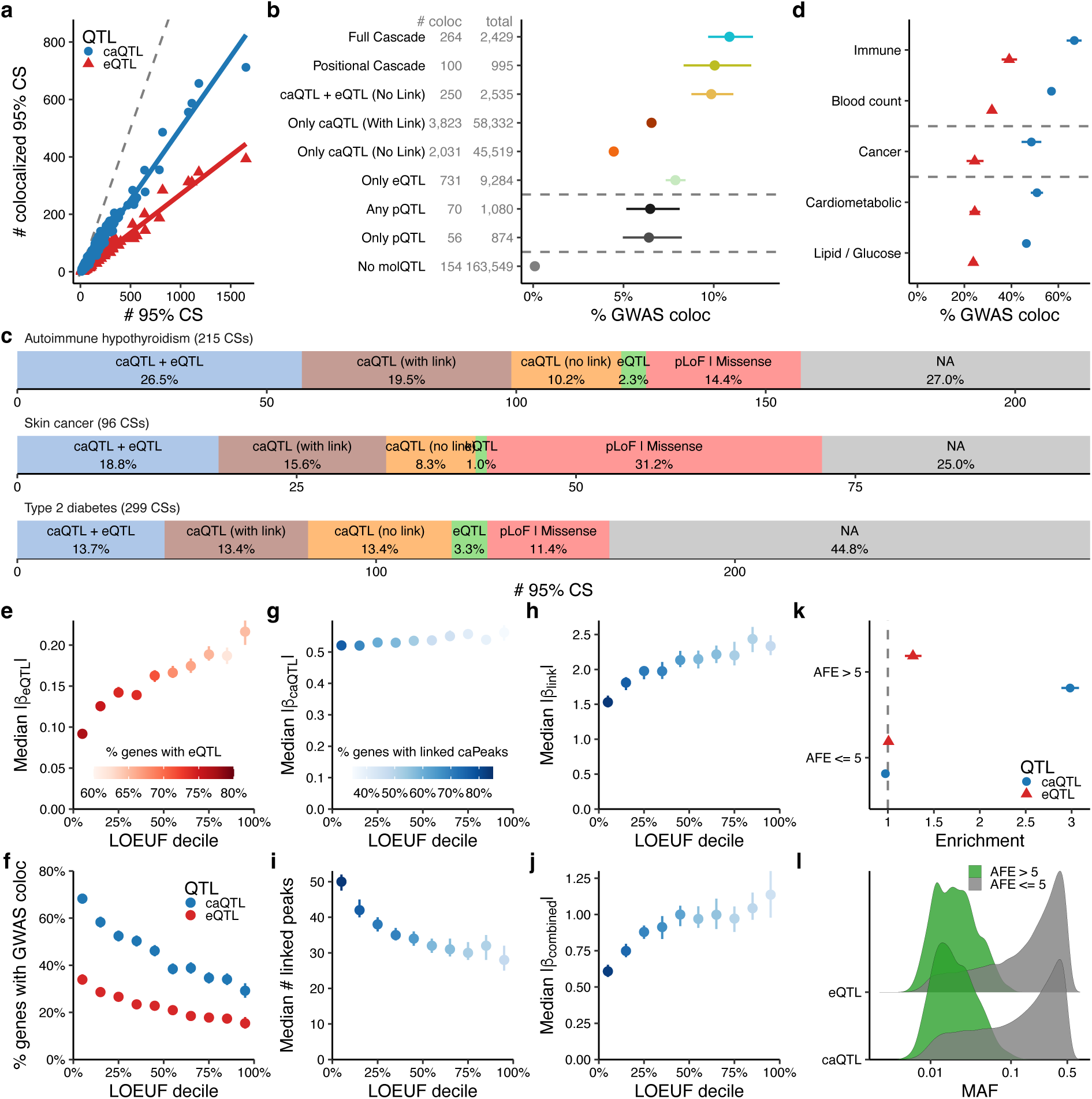
Colocalization between molQTL and GWAS variants reveals multi-layered regulatory buffering at constrained genes. **a.** Colocalization rates of 95% CSs from molQTL and 439 complex trait GWAS in FinnGen. The x-axis represents the number of 95% credible sets (CSs) for each trait. The y-axis represents the number of 95% CSs that are colocalized with either caQTL (blue) or eQTL (red) with PP.H4 > 0.5 (see **Methods**). Colored lines represent fitted slopes for caQTL and eQTL, whereas the dotted slope represents Y = X. **b**. Colocalization rates of fine-mapped molQTL variants (PIP > 0.5), stratified by QTL mechanism categories. “Any pQTL” and “Only pQTL” represent any fine-mapped pQTL (PIP > 0.5) variants and those not fine-mapped as caQTL or eQTL (PIP ≤ 0.5), respectively. Numbers of colocalized and total fine-mapped variants for each mechanism category are shown. Error bars represent binomial 95% confidence intervals. **c**. Proportion of 95% CSs classified by molQTL colocalization (caQTL & eQTL, only caQTL, and only eQTL) and presence of nonsynonymous coding variants (predicted loss-of-function [pLoF] or missense). Three example traits, autoimmune hypothyroidism, skin cancer, and type 2 diabetes, are shown to demonstrate the difference in molQTL colocalization rates across trait categories. **d**. Colocalization rates of 95% CSs from molQTL and GWAS from different trait categories. Error bars represent binomial 95% confidence intervals. **e–j**. The relationship between loss-of-function observed/expected upper bound fraction (LOEUF) deciles and regulatory layers. Panels show median absolute effect sizes for *cis*-eQTL (**e**), *cis*-caQTL (**g**), peak–gene links (**h**), and combined (*β*_combined_ = *β*_caQTL_ × *β*_link_) effect sizes (**j**), computed using variants with the strongest effect size for each gene (see **Methods**). Additional panels show the fraction of genes with GWAS colocalization through eQTL or linked peaks (**f**) and the median number of linked peaks per gene (**i**) are shown. Color represents the fraction of genes with either eQTL (blue) or linked peaks affected by caQTL (red) for each decile. The error bars represent 95% confidence intervals. **k**. Enrichment of fine-mapped caQTL or eQTL variants (PIP > 0.5) for Finnish allele frequency enrichment (AFE > 5). Enrichment is defined as a risk ratio of being fine-mapped and being AFE > 5 (**Methods**). Error bars represent 95% confidence intervals. **l**. The distribution of MAF for fine-mapped molQTL variants (PIP > 0.5) for Finnish-enriched (AFE > 5) and non-Finnish-enriched (AFE ≤ 5) variants.

To assess the functional importance of different regulatory mechanisms, we examined colocalization rates across CASCADE mechanistic categories (**Methods**). Variants with full cascades showed the highest GWAS colocalization rate at 11% (**Fig. 5b, Supplementary Table 24,25**), followed by positional cascades (10%) and caQTL + eQTL without links (9.8%). Notably, eQTL-only variants exhibited a relatively high colocalization rate (7.8%), suggesting that some disease variants operate through direct transcriptional regulation or chromatin states not fully captured in our current assays. Other partial regulatory mechanisms showed intermediate rates: caQTL-only with links (6.6%), and caQTL only without links (4.4%). When incorporating plasma *cis*-pQTLs from FinnGen (PIP > 0.5), pQTLs showed comparable colocalization rates to other molQTL-only categories (6.5% for any pQTL variants regardless of CASCADE status, 6.4% for pQTL-only variants without caQTL/eQTL effects; **Methods**), though with substantially fewer variants available. Variants with no detected molQTL effects (linkage disequilibrium [LD]-independent variants with max. PIP < 0.01 in both eQTL and caQTL, **Methods**) had minimal colocalization (0.05%), validating our regulatory classifications. These patterns suggest three mechanistic scenarios for disease variants: 1) those acting through regulatory cascades connecting chromatin to expression; 2) those affecting expression through mechanisms beyond chromatin accessibility changes detectable in baseline PBMCs; and 3) those affecting chromatin accessibility without detectable downstream effects on expression, potentially due to limited statistical power or context-specific expression regulation.

To illustrate how molecular QTLs elucidate disease mechanisms, we examined three exemplar diseases from different disease categories. Autoimmune hypothyroidism (AIHT), which has 215 fine-mapped 95% CSs in FinnGen, demonstrates a predominantly regulatory architecture: only two CSs harbor pLoF variants (0.9%) and 29 contain missense variants (13.5%), while the vast majority (58.5%) are colocalized with molQTLs—57 colocalized with both caQTL and eQTL (26.5%), 42 (19.5%) and 22 (10.2%) colocalized with caQTL only with and without link, respectively, and 5 colocalized with eQTL only (2.3%; **Fig. 5c, Supplementary Tables 26,27**). In contrast, skin cancer and type 2 diabetes show substantially lower molQTL colocalization rates (43.8% for both), particularly for caQTL and eQTL (18.8% and 13.7%, respectively; **Fig. 5c**), suggesting their genetic architecture involves regulatory mechanisms in other cell types beyond PBMCs. Together, 73% of CSs for AIHT can be attributed to either by nonsynonymous coding or molQTLs, generating multiple testable hypotheses to dissect immune disease mechanisms experimentally.

GWAS colocalization with the CASCADE framework revealed particularly striking examples of pleiotropic variants that influence AIHT risk through coordinated changes in chromatin accessibility and gene expression. Two completely linked variants upstream of *TICAM1* (rs10425559:A>G and rs10424978:C>A; *r*^2^ = 1.0) exemplify this mechanism. Both variants are protective against AIHT (OR = 0.95 for both, *P* = 7.3 × 10^-15^ and 8.4 × 10^-15^, PIP = 0.53 and 0.47, respectively) but increase risk for skin cancer (OR = 1.05 and 1.05, *P* = 7.5 × 10^-9^ and 9.2 × 10^-9^, PIP = 0.52 and 0.43; **Extended Data Fig. 7a**). Through a full cascade mechanism in CD16^+^ monocytes, these variants decrease chromatin accessibility (*β* = –0.40 for both, *P* = 7.8 × 10^-25^ and *P* = 8.0 × 10^-25^, PIP = 0.51 and 0.50) at a proximal peak (chr19:4837478-4837977; overlapping rs10424978 and located 3 bp away from rs10425559) and reduce *TICAM1* expression (*β* = –0.15 for both, *P* = 4.5 × 10^-33^ and 4.6 × 10^-33^, PIP = 0.50 and 0.49). *TICAM1* encodes an adaptor protein (also known as TRIF) that mediates TLR3 and TLR4 signaling to induce type I interferon production^50–52^. The decreased *TICAM1* expression in CD16^+^ monocytes likely reduces interferon-mediated autoimmune responses, providing protection against AIHT, while simultaneously impairing tumor immunosurveillance mechanisms, thereby increasing susceptibility to skin cancer.

Similarly, an intronic variant of *RNASET2*, rs3798307:G>A, demonstrates a positional cascade mechanism for *RNASET2* expression in monocytes with opposite effects on AIHT (OR = 0.94, *P* = 1.8 × 10^-16^) and skin cancer (OR = 1.06, *P* = 5.6 × 10^-9^; **Extended Data Fig. 7b**). In monocytes, rs3798307 increases *RNASET2* expression (*β* = 0.08, *P* = 1.9 × 10^-32^, PIP = 0.16) and overlaps with a chromatin accessibility peak (chr6:166950262-166950954) that is linked to *RNASET2* (*β* = 0.19, *P* = 5.0 × 10^-32^). *RNASET2* encodes a secreted ribonuclease that degrades extracellular RNA, a damage associated molecular pattern (DAMP) and potent immune activation signal, to limit inflammation^53^. Its increased expression could shift immune responses toward anti-inflammatory phenotypes, reducing thyroid inflammation while creating a tumor-permissive microenvironment.

The regulatory architecture of AIHT risk is further illustrated by rs13105678:C>A near *RHOH*, which demonstrates a full cascade mechanism in CD4^+^ T cells. This variant increases AIHT risk (OR = 1.04, *P* = 1.3 × 10^-8^, PIP = 0.70) by increasing *RHOH* expression (*β* = 0.02, *P* = 1.3 × 10^-8^, PIP = 0.99) and chromatin accessibility of an overlapping peak (chr4:40258141-40259058; *β* = 0.89, *P* = 1.8 × 10^-176^, PIP = 1.0) that is linked to *RHOH* (*β* = 0.15, *P* = 7.6 × 10^-57^; **Extended Data Fig. 7c**). *RHOH* encodes a GTPase-deficient Rho family protein that recruits ZAP70 kinase to the T cell receptor complex^54^. Intriguingly, we identified a Finnish-enriched missense variant in *ZAP70* itself (rs145955907:C>T, p.Thr155Met, MAF = 2%) that independently increases AIHT risk (OR = 1.26, *P* = 2.3 × 10^-23^, PIP = 0.25)^46^. While the interaction between these variants was sub-significant (OR = 1.04, *P* = 0.1), the convergence of regulatory and coding variation on T cell receptor signaling components highlights how multiple genetic mechanisms can converge on shared pathways.

These disease-specific patterns reflect broader trends across disease categories. Immune-related traits consistently showed the highest colocalization rates for both caQTLs and eQTLs (66.8% and 39.1%, respectively), followed by cancer (48.5% and 24.4%) and cardiometabolic traits (50.9% and 24.4%; **Fig. 5d, Supplementary Table 28**). Similar patterns emerged for quantitative traits, with blood counts showing higher colocalization rates (57.1% and 31.7% for caQTLs and eQTLs, respectively) than lipid and glucose traits (46.4% and 23.8%; **Fig. 5d**). These systematic differences underscore the importance of matching molecular QTL data to disease-relevant cell types.

The utility of molQTLs in PBMCs extends beyond immune diseases to conditions where circulating immune cells influence pathology in other tissues. An intergenic variant rs6979218:C>G exemplifies this principle by its increased risk for Alzheimer’s disease (OR = 1.08, *P* = 1.3 × 10^-15^ in the latest meta-analysis^55^; **Extended Data Fig. 8a**) and a full cascade mechanism in monocytes. This variant decreases *PILRB* expression (*β* = –0.07, *P* = 4.1 × 10^-25^, PIP = 1.0) and chromatin accessibility of its overlapping peak (chr7:100295229-100295728, *β* = –1.3, *P* = 1.5 × 10^-303^, PIP = 0.02), which is linked to *PILRB* (*β* = 0.90, *P* = 1.1 × 10^-66^). *PILRB* encodes a paired immunoglobulin-like type 2 receptor that activates myeloid cells by signaling through DAP12 (encoded by *TYROBP*)^56^. DAP12 deficiency in humans has been linked to early-onset dementia^57^. Notably, rs1244787406:T>G (MAF = 0.27%), which tags a 5.2 kb Finnish-enriched *TYROBP* deletion recently identified as an Alzheimer’s disease risk factor^58^, significantly decreases *TYROBP* expression in monocytes (*β* = –0.51, *P* = 3.3 × 10^-20^, **Extended Data Fig. 8b, Methods**), despite the presence of only six heterozygous carriers in our multiome samples. Given the shared developmental origin and functional similarities between peripheral monocytes and brain microglia, decreased *PILRB* expression or DAP12 levels could enhance neuroinflammation and impair amyloid-β clearance.

Similarly, the *LTBR* locus provides another compelling example of how molQTLs in PBMCs can reveal shared regulatory mechanisms across anatomically distinct inflammatory conditions. A 5’ UTR variant of *LTBR*, rs10849448:A>G, is protective against chronic diseases of tonsils and adenoids (OR = 0.90, *P* = 1.0 × 10^-41^, PIP = 1.0) but is risk for acute appendicitis (OR = 1.09, *P* = 1.0 × 10^-23^, PIP = 0.99; **Extended Data Fig. 8c**). Through a positional cascade mechanism in monocytes, rs10849448 decreases *LTBR* expression (*β* = –0.05, *P* = 1.4 × 10^-11^, PIP = 0.75), and overlaps with a chromatin accessibility peak (chr12:6383829-6384328) that is linked to *LTBR* (*β* = –0.10, *P* = 0.02). LTBR mediates lymphotoxin signaling essential for lymphoid organogenesis, germinal center formation, and dendritic cell homeostasis^59,60^. Decreased *LTBR* expression could reduce chronic lymphoid hyperplasia in tonsils while compromising acute antimicrobial responses in the appendix, illustrating how the same regulatory variant can have opposing effects depending on the tissue context and inflammatory stimulus.

### Quantifying the regulatory architecture of disease heritability

To quantify the proportion of disease heritability mediated by molecular traits, we applied mediated expression score regression (MESC)^24^ using *cis*-eQTLs and *cis*-caQTLs from PBMC cell types as well as plasma *cis*-pQTLs from FinnGen (**Methods**). For immune diseases, *cis*-caQTLs mediated 79 ± 4% of disease heritability, with *cis*-eQTLs mediating 25 ± 2% and *cis*-pQTLs 4 ± 0.4% (**Extended Data Fig. 9a, Supplementary Table 29**). Notably, the combined contribution of all three QTL types did not exceed that of caQTLs alone, indicating that caQTLs are the primary mediator through which regulatory variation manifests as gene expression and protein abundance differences. For cancer and cardiometabolic diseases, caQTLs mediated 45 ± 3% and 34 ± 2% of disease heritability, respectively, highlighting the importance of quantifying disease-relevant cell types.

To partition heritability by chromatin accessibility peaks, we applied stratified LD score regression (S-LDSC)^61^, which captures heritability enrichment of genomic annotations regardless of mediation (**Methods**). All chromatin accessibility peaks captured 65 ± 5% of immune disease heritability, 35 ± 4% for cancer, 12 ± 2% for cardiometabolic diseases (**Extended Data Fig. 9b, Supplementary Table 30**), compared to 79%, 45%, and 34% from MESC estimates for caQTL-mediated heritability, respectively. When stratified by peak–gene link status, peaks linked to genes (CA-Link+) showed substantially higher enrichment (21-fold) compared to peaks without links (CA-Link–; 9-fold) for immune diseases (**Extended Data Fig. 9c**). This enrichment difference diminished for non-immune diseases, consistent with the cell-type specificity of immune regulatory effects. Together, these analyses establish chromatin accessibility as a primary mediator of disease heritability, particularly for immune diseases where disease-relevant cell types were directly profiled. Nevertheless, eQTL-only variants retain non-negligible disease colocalization (7.8%; **Fig. 5b**), indicating regulatory mechanisms independent of detectable chromatin accessibility.

### Multi-layered regulatory buffering at constrained genes

Previous studies have identified an apparent paradox in human genetics: evolutionarily constrained genes (those for which natural selection does not tolerate significant coding mutation) are enriched for proximity to GWAS-discovered disease associations, yet show depletion of detectable *cis*-eQTLs^8,21,22,35^. To investigate this paradox, we systematically characterized the regulatory architecture of 15,976 protein-coding genes expressed in PBMCs across the constraint spectrum, examining *cis*-eQTLs, *cis*-caQTLs, and peak–gene links using loss-of-function observed/expected upper bound fraction (LOEUF) deciles from gnomAD^62,63^ v4.1 (**Methods**).

We found a striking gradient across the constraint spectrum: genes in the most constrained decile (lowest LOEUF decile, hereafter referred to as “constrained genes”) were more likely to harbor *cis*-eQTLs but with significantly smaller effect sizes compared to genes in the least constrained decile (highest LOEUF decile, hereafter referred to as “unconstrained genes”; median |*β*_eQTL_| = 0.09 vs 0.22; Spearman’s *ρ* = 0.20, *P* = 1.7 × 10^-2^^28^ for correlation between LOEUF and |*β*_eQTL_|; **Fig. 5e, Extended Data Fig. 9d**). Consistent with previous findings^35,62,64^, constrained genes showed higher mean expression (Spearman’s *ρ* = –0.33, *P* = 2.0 × 10^-386^) with lower variance (Spearman’s *ρ* = 0.07, *P* = 2.7 × 10^-13^ for correlation between LOEUF and expression variance residuals corrected by mean expression); however, the eQTL effect size gradient remained robust even among expression-matched highly-expressed genes (top 20%; Spearman’s *ρ* = 0.23, *P* = 7.6 × 10^-18^), confirming that our observed pattern is not driven by expression-level differences (**Extended Data Fig. 9e–g, Methods**). Despite these weaker individual effects, *cis*-eQTLs at highly constrained genes were more likely to colocalize with GWAS traits—34% of constrained genes showed colocalization compared to only 15% of unconstrained genes (**Fig. 5f**). This enrichment suggests that even modest expression changes in highly constrained genes, which selection has fine-tuned to operate within narrow functional ranges, can disrupt critical molecular mechanisms and contribute to disease pathogenesis. The apparent discrepancy with earlier findings^8,21,22,35^ is primarily power-based: while the largest eQTLs detectable in small, bulk RNA studies indeed appear enriched at unconstrained and non-disease relevant genes, increased power reveals that constrained genes were actually more likely to harbor eQTLs, albeit with much smaller average effects.

For example, we found strong *cis*-eQTLs at constrained genes *GRAMD1B* and *FAS* that colocalize with chronic lymphocytic leukaemia (CLL) risk. An intron variant of *GRAMD1B*, rs35923643:A>G, substantially increases *GRAMD1B* expression in B cells (*β* = 0.66, *P* = 3.7 × 10^-59^), while increasing CLL risk (OR = 1.58, *P* = 5.0 × 10^-71^ in the FinnGen+MVP+UKB meta-analysis; **Extended Data Fig. 10a, b**). *GRAMD1B* encodes a cholesterol transfer protein that regulates lipid homeostasis at ER-plasma membrane contact sites^65^, and rs35923643 is shown to be associated with increased transitional B cells and decreased levels of proteins involved in CLL cell survival (SIRT2 and ADA)^66^, potentially promoting the accumulation of premalignant B cells characteristic of CLL. Similarly, an intron variant of *FAS*, rs2147420:A>G increases *FAS* expression across immune cells, with the strongest effect in CD16^+^ monocytes (*β* = 0.53, *P* = 4.9 × 10^-89^) and a moderate effect in B cells (*β* = 0.20, *P* = 1.2 × 10^-29^), while increasing CLL risk (OR = 1.27, *P* = 6.9 × 10^-28^; **Extended Data Fig. 10c,d**). *FAS* encodes a death receptor that triggers apoptosis upon engagement by FAS ligand, and paradoxically, its increased expression in CLL may reflect compensatory upregulation in response to apoptosis resistance mechanisms or could promote survival signals through non-canonical FAS pathways that have been implicated in B cell proliferation.

To dissect how selection shapes regulatory architecture across molecular layers, we examined *cis*-caQTLs mapped to 70,803 peaks linked to 9,984 expressed genes. While constrained genes were more likely to harbor linked peaks affected by caQTLs than unconstrained genes (76% vs. 36%), the effect sizes of these caQTLs showed only a weak relationship with constraint (median |*β*_caQTL_| = 0.52 vs. 0.56; Spearman’s *ρ* = 0.02, *P* = 3.1 × 10^-10^; **Fig. 5g, Extended Data Fig. 9d**), in stark contrast to the strong correlation observed for eQTLs. This difference between eQTL and caQTL patterns persists when examining MAF-normalized effect sizes, indicating that this pattern is not driven by differences in statistical power (**Extended Data Fig. 9h,i**). In parallel, analysis of peak–gene links revealed that constrained genes had more peak–gene links but with smaller effect sizes compared to unconstrained genes (median number of linked peaks = 50 vs. 28; median |*β*_link_| = 1.5 vs. 2.3; Spearman’s *ρ* = 0.16, *P* = 5.8 × 10^-59^; **Fig. 5h,i**). The relationship between peak and gene constraint showed distinct patterns: constrained peaks preferentially linked to constrained genes (median genomic non-coding constraint of haploinsufficient variation [Gnocchi]^63^ = 5.2 vs. 4.8 for the most constrained peak per gene), yet the strongest peak per gene showed no correlation with gene constraint (**Extended Data Fig. 9j**). Stratifying all peak–gene links by both peak and gene constraint revealed that unconstrained peaks (–4 < Gnocchi < 4) showed stronger regulatory effects than constrained peaks (Gnocchi > 4) peaks, with both peak constraint categories showing similar gene constraint (LOEUF)-dependent increases (**Extended Data Fig. 9k**), suggesting that constrained peaks are systematically weakened to maintain essential gene expression. To approximate *cis*-eQTL effects, we integrated these two regulatory layers by calculating the combined effect of genetic variants on gene expression through chromatin accessibility (*β*_combined_ = *β*_caQTL_ × *β*_link_) and observed striking attenuation at constrained genes compared to unconstrained genes (median |*β*_combined_| = 0.61 vs. 1.1; Spearman’s *ρ* = 0.18, *P* = 8.4 × 10^-71^; **Fig. 5j**), recapitulating the constraint gradient we observed for empirical *cis*-eQTLs (**Fig. 5e**). This reveals a hierarchical buffering architecture where genetic variants alter chromatin accessibility with largely uniform effect sizes across the constraint spectrum, but the transmission of these changes to gene expression is progressively attenuated at constrained genes through systematically weaker peak–gene links.

Remarkably, caQTLs affecting peaks linked to constrained genes showed even higher GWAS colocalization rates than eQTLs—68% of constrained genes with linked caQTLs colocalized with GWAS traits compared to 29% of unconstrained genes (**Fig. 5f**). This enhanced colocalization with caQTLs suggests that many disease-associated variants operate primarily through chromatin accessibility changes that are buffered before reaching gene expression, providing a complementary path for disease risk that circumvents the strong selective constraints on expression variation at essential genes.

These findings provide mechanistic insights into the GWAS–eQTL colocalization paradox through a model of multi-stage regulatory buffering^67^, extending theoretical predictions that natural selection produces observed differences between GWAS and eQTL studies^22^. At constrained genes where even modest expression changes would have large phenotypic consequences, evolution has implemented a sophisticated buffering system that operates differentially across molecular layers. Genetic variants can alter chromatin states with normal effect sizes, allowing tolerable variation at this regulatory tier, but buffering is enforced through a larger number of attenuated peak–gene links that create a regulatory bottleneck preventing chromatin changes from strongly impacting expression. This architecture aligns with the proposed selection model where variants with large expression effects at constrained genes are purged due to their deleterious phenotypic impact, leaving only small-effect variants that require large sample sizes to detect^22^. The differential buffering across molecular layers explains why caQTLs remain readily detectable at constrained genes—they operate upstream of the main buffering checkpoint at the chromatin-to-expression interface—and why they show even higher GWAS colocalization rates than eQTLs, as they capture disease-relevant regulatory variation before it is attenuated by peak–gene buffering.

### Molecular mechanisms of complex diseases

Among 119,094 fine-mapped molQTL variants (PIP > 0.5), 8,097 variants are Finnish-enriched (allele frequency enrichment [AFE] > 5x in Finnish vs non-Finnish non-Estonian Europeans; **Supplementary Table 31**, **Methods**). Of these, 121 showed full cascade and 42 showed positional cascade mechanisms. Notably, these Finnish-enriched variants cannot be properly characterized in non-Finnish populations: only 78 Finnish-enriched variants (1%) were fine-mapped (PIP > 0.5) in the eQTL Catalogue (R7)^7^.

We previously demonstrated that Finnish-enriched variants, particularly those stochastically boosted through bottleneck events, are enriched among high-PIP complex trait variants^68^. Extending this to molQTLs, we observed that fine-mapped caQTL variants (PIP > 0.5) are enriched for Finnish-enriched (AFE > 5×) variants (RR = 2.98), whereas fine-mapped eQTL variants show attenuated enrichment (RR = 1.27; **Fig. 5k, Supplementary Table 32**). Of note, the MAF distributions of fine-mapped molQTL variants (PIP > 0.5) are similar between caQTLs and eQTLs for both Finnish-enriched (AFE > 5×) and non-enriched (AFE ≤ 5×) variants (**Fig. 5l**), indicating that the observed enrichment pattern is not simply driven by differences in statistical power. Given that Finnish-enriched variants escape negative selection through population bottlenecks and tend to be disease-causing and deleterious^34,68^, this divergent enrichment pattern provides compelling support for our multi-layered buffering model. The enrichment of Finnish-enriched variants among caQTLs demonstrates that deleterious regulatory variation accumulates at the chromatin level where it is buffered from expression. This creates a reservoir of disease-relevant variation that would otherwise be purged if it directly impacted gene expression levels.

Here, we highlight two illustrative examples of Finnish-enriched variants with strong evidence for causality in immune-mediated diseases as elucidated by our molecular QTLs. First, an intronic variant of *TNRC18*, rs748670681:C>T, is substantially enriched in the Finnish population (MAF = 3.6% in Finnish, AFE = 114x) and showed varying effects across multiple autoimmune diseases (**Fig. 6a,b**)^34^, with a strong risk for IBD (odds ratio [OR] = 2.2, *P* = 7.9 × 10^-157^, PIP = 1.0) but protective against AIHT (OR = 0.89, *P* = 4.3 × 10^-9^, PIP = 0.91). Using our molecular QTLs, we identified that rs748670681 is a positional cascade variant—*i.e.*, it showed a strong *cis*-eQTL for *TNRC18* (*β* = –0.46, *P* = 9.0 × 10^-128^, PIP = 1.0 in CD8^+^ T cells), with varying effect sizes among lymphoid cell types (**Fig. 6c,d**), overlapped with a peak (chr7:5397119-5397618), and the overlapping peak is significantly associated with *TNRC18* expression in CD8^+^ T cells (*β* = –0.33, *P* = 1.5 × 10^-62^; **Fig. 6e,f**), but rs748670681 was not a caQTL for the overlapping peak within the same cell type.

**Fig. 6:**
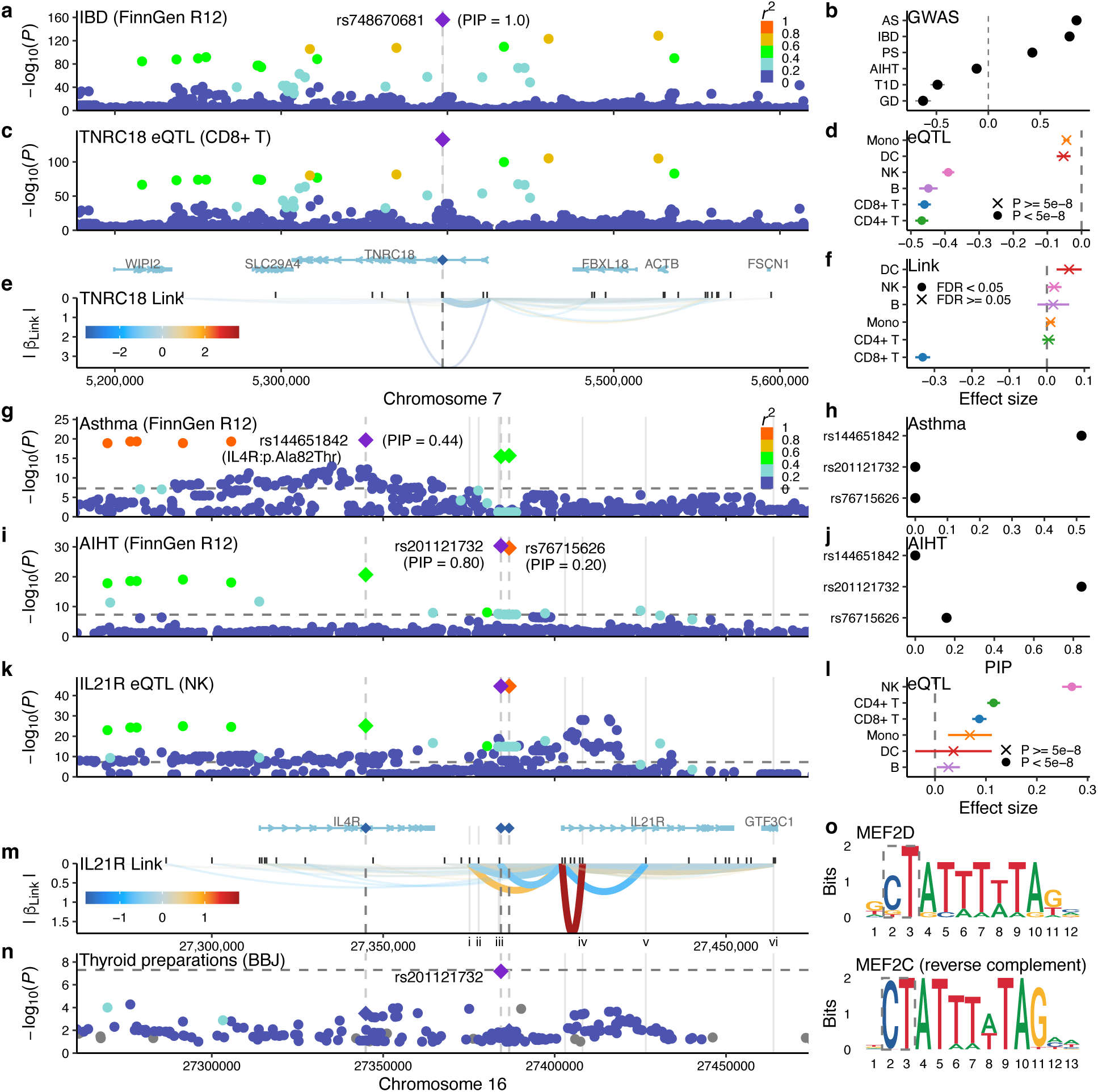
Illustrative vignettes of Finnish-enriched disease-causing variants explained by molQTL mechanisms. **a.** LocusZoom plot of inflammatory bowel disease (IBD) in FinnGen for the *TNRC18* locus. A lead variant, rs748670681, is shown as a diamond. Color represents *r*^2^ to the lead variant. **b**. Forest plot of rs748670681 effect sizes across autoimmune diseases in FinnGen. AS, ankylosing spondylitis. IBD, inflammatory bowel disease. PS, psoriasis. AIHT, autoimmune hypothyroidism. T1D, type I diabetes. GD, Grave’s disease. **c**. LocusZoom plot of *cis*-eQTL effect for *TNRC18* expression in CD8^+^ T cells. **d**. Forest plot of rs748670681 effect sizes for *TNRC18* expression across cell types. **e**. Peak–gene links between chromatin accessibility (CA) peaks and *TNRC18* expression across cell types. Each curved line represents a link, where the bold line represents the peak (chr7:5397119-5397618) involved in the positional cascade mechanism. Color represents effect sizes of peak–gene links, whereas Y-axis represents the corresponding absolute value. Marginal rugs at the top represent CA peak locations. **f**. Forest plot of peak–gene links (chr7:5397119-5397618 and *TNRC18*) across cell types. **g**. LocusZoom plot of asthma in FinnGen for the *IL4R/IL21R* locus. **h**. PIPs of rs144651842, rs201121732, and rs76715626 in asthma. **i**. LocusZoom plot of AIHT for the *IL4R/IL21R* locus. **j**. PIPs of rs144651842, rs201121732, and rs76715626 in AIHT. **k**. LocusZoom plot of *cis*-eQTL effect for *IL21R* expression in NK cells. **l**. Forest plot of rs201121732 effect sizes for *IL21R* expression across cell types. **m**. Peak–gene links between CA peaks and *IL21R* expression across cell types. **n**. LocusZoom plot of thyroid preparations (ATC code: H03A) medication in BioBank Japan (BBJ) for the IL4R/IL21R locus. **o**. Sequence logos for MEF2D and MEF2C (reverse complement) transcription factors. The dotted rectangle represents the location of the rs201121732 variant.

In a companion paper^69^, we developed a novel base-editing and phenotyping platform called Sensitive Transcriptomics And Genotyping by sequencing (STAG-seq), which validated that the minor allele for rs748670681 is associated with *TNRC18* down-regulation and provided deeper mechanistic insight showing the strongest effect in Th1 among primary immune cells. In Th1, we demonstrated that rs748670681 reactivated endogenous retroviral elements, up-regulated interferon-stimulated genes, and suppressed cell-cycle transcripts, shifting the population toward a memory-like inflammatory state. These coordinated effects delineate a causal path from a non-coding variant to heightened autoimmune susceptibility.

Furthermore, we identified a Finnish-enriched haplotype associated with immune-related diseases that exhibits two partially correlated but distinct putative causal variants. On 16p12.1, a missense variant of *IL4R*, rs144651842:G>A (p.Ala82Thr, MAF = 7.7%, AFE = 119x) is significantly associated with asthma (OR = 0.88, *P* = 2.6 × 10^-24^, PIP = 0.44; **Fig. 6g,h**). In addition, two distinct intergenic variants between *IL4R* and *IL21R*, rs201121732:C>CT and rs76715626:T>C, are significantly associated with AIHT (OR = 1.19 for both, *P* = 2.2 × 10^-34^ and 8.5 × 10^-34^, PIP = 0.80 and 0.20, respectively; **Fig. 6i,j**). These two variants are in near-complete LD (*r*^2^ = 0.98), and modestly correlated with the asthma-associated missense variant rs144651842:G>A (*r*^2^ = 0.60). By leveraging our molecular QTLs, these intergenic variants are *cis*-eQTL for *IL21R* in T and NK cells (*β* = 0.27, *P* = 1.4 × 10^-44^, PIP = 0.5 in NK cells; **Fig. 6k, l**). They are *cis*-caQTL for 16 nearby peaks, including six peaks that showed significant association with *IL21R* expression in NK and CD4^+^ T cells, such as the most proximal peak (chr16:27383475-27384329, located 12 bp away from rs201121732; **Fig. 6m**). Furthermore, we found that only rs201121732, not rs76715626, is sub-significantly associated with thyroid preparations (ATC code: H03A) medication (OR = 1.45, *P* = 6.5 × 10^-8^; **Fig. 6n**) in BioBank Japan (BBJ)^70^, where the two variants are no longer in LD in the Japanese population (*r*^2^ = 0.2). Fine-mapping of *IL21R cis*-eQTL in bulk whole blood samples from the Japanese population^71^ also pinpoints rs201121732 (*β* = 0.21; *P* = 4.7 × 10^-8^; PIP = 1.0), recapitulating our observations. Finally, motif analysis identified that rs201121732 increases affinity to MEF2 transcription factors which target IL21R (**Fig. 6o, Methods**).

Together, these examples demonstrate how population-enriched variants illuminate universal regulatory mechanisms. The *TNRC18* variant exemplifies a mechanistic cascade where chromatin changes transmit to gene expression, with functional validation revealing downstream consequences such as retroviral reactivation and inflammatory memory states. The *IL4R/IL21R* haplotype demonstrates how the integration of population-scale molQTLs and cross-population fine-mapping pinpoints distinct causal variants on the same Finnish-enriched haplotype. The Finnish population’s unique genetic architecture enabled discovery of these regulatory variants that would remain undetected in other populations. More broadly, our population-scale molQTLs identified regulatory mechanisms for over 100,000 variants, including both common variants shared across populations and Finnish-enriched variants, with the molecular cascades from chromatin accessibility to gene expression. This represents fundamental biology applicable across diverse populations, establishing a paradigm for leveraging population-scale molQTLs with base-editing validation to comprehensively map the regulatory landscape of complex diseases.

## Discussion

Our population-scale single-nucleus multiome atlas of 1,108 Finnish individuals generated paired gene expression and chromatin accessibility profiles from 10 million PBMC nuclei, establishing the most comprehensive immune regulatory map to date. We identified 20,829 genes and 210,584 peaks (76% and 71% of tested features, respectively) under *cis*-genetic regulation, fine-mapped 16,477 *cis*-eQTL and 109,163 *cis*-caQTL variants (PIP > 0.5), and mapped 496,488 enhancer–gene links connecting 112,032 peaks to 12,445 genes. This foundational resource enabled comprehensive dissection of regulatory cascades through our CASCADE framework, which systematically classified 119,094 fine-mapped molQTL variants into distinct mechanistic categories. We identified complete regulatory pathways for 3,424 fine-mapped variants (2,429 full cascade and 995 positional cascade variants), with an additional 70,151 variants connecting to target genes through enhancer–gene links or eQTL effects, demonstrating our capacity to precisely map the multi-layered regulatory architecture underlying genetic variation.

Colocalization with 439 GWAS traits in FinnGen revealed that molecular regulatory mechanisms predict disease relevance. Fine-mapped molQTL variants with regulatory cascades showed 2-fold higher colocalization rates (9.8–11%) than those affecting only chromatin (4.4–6.6%), establishing a clear hierarchy where mechanistic cascade correlates with disease association. This pattern was most evident for immune traits, which showed substantially higher colocalization rates (66.8% for caQTLs, 39.1% for eQTLs) than other trait categories, reflecting the relevance of PBMCs to immune disease biology. Moreover, the utility of our molQTL resource in PBMCs extends beyond immune diseases to conditions where circulating immune cells influence pathology in distant tissues, as exemplified by the *PILRB* variant affecting microglial-related gene linked to Alzheimer’s disease and the *LTBR* variant associated with inflammatory conditions across anatomically distinct sites. Across all traits, caQTLs showed nearly twice the GWAS colocalization rate of eQTLs (51.7% vs 27.4%), indicating that chromatin accessibility captures disease-relevant regulatory variation at its point of origin, upstream of transcriptional regulation. These patterns delineate three distinct mechanistic scenarios for disease variants: i) those acting through complete regulatory cascades from chromatin to expression, ii) those affecting expression through mechanisms beyond detectable chromatin accessibility changes in baseline PBMCs, and iii) those affecting chromatin without downstream expression effects, potentially due to limited statistical power or context-specific regulation. This hierarchical organization of regulatory effects becomes particularly important when considering how evolutionary constraints shape the transmission of genetic effects across molecular layers.

Analysis of constrained genes revealed a multi-layered regulatory buffering architecture that reconciles the long-standing GWAS–eQTL colocalization paradox, where disease-associated variants preferentially target constrained genes yet show depletion of detectable eQTL colocalization in prior studies^8,21,22,35^. Our population-scale molQTLs enabled us to detect that constrained genes actually harbor more *cis*-eQTLs and show higher GWAS–eQTL colocalization rates than unconstrained genes (34% vs 15%), despite having systematically smaller eQTL effect sizes, revealing that the apparent discrepancy with earlier findings was primarily due to insufficient power to detect these small-effect eQTLs. The pattern of small eQTL effects yet high disease colocalization is explained by differential buffering across molecular layers: *cis*-caQTLs affecting peaks linked to constrained genes showed comparable effect sizes to those for unconstrained genes, but constrained genes exhibited more numerous yet weaker peak–gene links that attenuate transmission of chromatin changes to gene expression, implementing regulatory buffering that prevents chromatin changes from strongly impacting expression of essential genes. Consequently, caQTLs affecting peaks linked to constrained genes showed even higher GWAS colocalization rate than eQTLs (68% vs 34%), as they capture disease-relevant regulatory variation upstream of the main buffering checkpoint at the enhancer–gene links, providing a complementary path for disease risk that circumvents the strong selective constraints on expression variation at essential genes.

Finnish-enriched variants provide unique insights into disease mechanisms that remain uncharacterized in other populations^34,68^. These variants are enriched among caQTLs but attenuated among eQTLs (RR = 2.98 and 1.27, respectively), despite similar MAF distributions, suggesting that deleterious regulatory variation preferentially manifests at the chromatin level before being buffered at transcription. The *TNRC18* locus exemplifies how founder populations enable mechanistic discovery—an intron variant (rs748670681) with 114-fold enrichment in the Finnish population shows opposite effects on IBD and AIHT, and our integration with STAG-Seq^69^ revealed it reactivates endogenous retroviral elements and shifts Th1 cells toward inflammatory memory states. Prioritizing such molecular mechanisms for experiments would be intractable without the natural genetic variants provided by population bottlenecks. Only 1% of Finnish-enriched molQTL variants are fine-mapped in non-Finnish datasets, underscoring that comprehensive architecture requires diverse populations. The regulatory architecture and molecular mechanisms we establish represent fundamental biology applicable across human populations. The Finnish population’s unique genetic history thus provides an invaluable window into universal mechanisms of immune regulation.

Our work demonstrates a powerful paradigm integrating population-scale single-cell molQTLs with precise experimental validation. While molQTL mapping identifies regulatory variants and predicts their mechanisms through our CASCADE classification, high-throughput base-editing platforms like STAG-Seq^69^ provide definitive functional evidence and reveal deeper mechanistic insights. For *TNRC18*, population-scale single-cell molQTL identified the putative causal variant and cell type, while base-editing uncovered specific downstream consequences including retroviral reactivation. This synergy enables population-scale molQTL discovery followed by mechanistic validation of disease-causing genetic variants, creating a complete pipeline from causal variants to molecular functions (V2F).

Looking forward, we envision an integrated framework for human genetics that systematically combines population-scale single-cell molQTLs with experimental perturbation across diverse populations, cell types, and physiological states. Our molQTL discovery for baseline PBMCs approaches saturation for common variants in steady-state immune cells, yet this represents only a fraction of the full regulatory landscape. Profiling stimulus-responsive regulatory programs, where genetic effects emerge only after immune stimulation, will be essential for capturing additional disease-relevant biology, as the emerging field of single-cell dynamic QTL mapping demonstrates that substantial fractions of disease-associated variants show context-dependent effects^14,18,72^. Similarly, tissue-resident immune cells and disease-specific cell states beyond PBMCs harbor distinct regulatory architectures critical for understanding tissue-specific autoimmunity and inflammatory diseases. The sparsity inherent to single-cell profiling, while partially addressed by our large sample size and deeper profiling per individual, remains a fundamental challenge that novel statistical and experimental methods may overcome.

Most importantly, systematic base editor screening across multiple cell types and states offers a path to functionally validate causality at scale. Unlike GWAS and molQTL studies, which are inevitably constrained by LD and lack of functional validation, base editing provides unambiguous evidence though direct nucleotide manipulation and phenotypic measurement. High-throughput platforms combining base editing with transcriptional and immunophenotypic readouts in primary human cells will bridge the gap between genetic association and biological mechanism. The integration of population-scale single-cell molQTL with systematic base editor screens has the potential to transform human genetics into a complete discovery engine—generating hypotheses at scale through molQTLs and validating mechanisms through experimental perturbation—thereby realizing the promise of GWAS to illuminate the biological basis of complex diseases and enable rational therapeutic development.

## Data availability

The FinnGen single-nucleus caQTL and eQTL summary statistics are currently subject to embargo according to the FinnGen consortium agreement. They are available upon request (https://www.finngen.fi/en/access_results) and are being prepared for public release by Q4 2026 or at the time of publication. The interactive CASCADE browser will be publicly available upon public release. The FinnGen GWAS summary statistics (R12) are available at https://r12.finngen.fi/. The KANTA lab value GWAS summary statistics are available at https://labvalues.finngen.fi/. The FinnGen (R12) + MVP + UKBB meta-analysis results are available at https://mvp-ukbb.finngen.fi/.

## Code availability

Our single-cell processing pipelines and analysis workflows will be made available at https://github.com/FINNGEN/singlecell-pipeline. The code to generate the figures will be made available at https://github.com/mkanai/finngen-multiome-flagship. The CASCADE framework will be made available at https://github.com/mkanai/cascade. The fasthurdle package for efficient hurdle model computation is available at https://github.com/mkanai/fasthurdle. The ldcov package for efficient covariate-adjusted LD computation will be made available at https://github.com/mkanai/ldcov. FinnGen’s standard analysis workflows are available as follows: GWAS, https://github.com/FINNGEN/regenie-pipelines; Fine-mapping, https://github.com/FINNGEN/finemapping-pipeline; Colocalization, https://github.com/FINNGEN/coloc.susie.direct; KANTA preprocessing & QC: https://github.com/FINNGEN/kanta_lab_preprocessing.

## Supporting information

Supplementary Information - FinnGen banner

## Data Availability

https://r12.finngen.fi/

https://labvalues.finngen.fi/

https://mvp-ukbb.finngen.fi/

## Acknowledgements

This work was supported by funding from the Klarman Cell Observatory and sundry funds from the Department of Molecular Biology at MGH to R.J.X. M.Kanai was supported by the Masason Foundation. We thank the blood donors and the staff of the Finnish Red Cross Blood Service for their help in sample collection. We want to acknowledge the participants and investigators of the FinnGen study. The FinnGen project is funded by two grants from Business Finland (HUS 4685/31/2016 and UH 4386/31/2016) and the following industry partners: AbbVie Inc., AstraZeneca UK Ltd, Biogen MA Inc., Bristol Myers Squibb Inc. (and Celgene Corporation & Celgene International II Sàrl), Genentech Inc., Merck Sharp & Dohme LCC, Pfizer Inc., GlaxoSmithKline Intellectual Property Development Ltd., Sanofi US Services Inc., Maze Therapeutics Inc., Johnson&Johnson Innovative Medicine Inc., Novartis AG, Boehringer Ingelheim International GmbH and Bayer AG. Following biobanks are acknowledged for delivering biobank samples to FinnGen: Auria Biobank (www.auria.fi/biopankki), THL Biobank (www.thl.fi/biobank), Helsinki Biobank (www.helsinginbiopankki.fi), Biobank Borealis of Northern Finland (https://www.ppshp.fi/Tutkimus-ja-opetus/Biopankki/Pages/Biobank-Borealis-briefly-in-English.aspx), Finnish Clinical Biobank Tampere (www.tays.fi/en-US/Research_and_development/Finnish_Clinical_Biobank_Tampere), Biobank of Eastern Finland (www.ita-suomenbiopankki.fi/en), Central Finland Biobank (www.ksshp.fi/fi-FI/Potilaalle/Biopankki), Finnish Red Cross Blood Service Biobank (www.veripalvelu.fi/verenluovutus/biopankkitoiminta), Terveystalo Biobank (www.terveystalo.com/fi/Yritystietoa/Terveystalo-Biopankki/Biopankki/) and Arctic Biobank (https://www.oulu.fi/en/university/faculties-and-units/faculty-medicine/northern-finland-birth-cohorts-and-arctic-biobank). All Finnish Biobanks are members of BBMRI.fi infrastructure (https://www.bbmri-eric.eu/national-nodes/finland/). Finnish Biobank Cooperative-FINBB (https://finbb.fi/) is the coordinator of BBMRI-ERIC operations in Finland. The Finnish biobank data can be accessed through the Fingenious® services (https://site.fingenious.fi/en/) managed by FINBB. This work is also supported by the FinnGen project for the costs of sample collection and single-cell sequencing.

## Methods

### Study cohort

FinnGen is a public-private partnership project combining genotype data from Finnish biobanks and digital health records from Finnish health registries^34^. Genotype and phenotype data in this study is based on the Data Freeze 12 (October 2023) which contains 500,348 individuals of Finnish ancestry. FinnGen participants provided informed consent as described in **Supplementary Note**. Detailed characteristics of the cohort are described elsewhere^34^.

Samples were primarily genotyped using the FinnGen ThermoFisher Axiom custom array. The samples from legacy cohorts have previously been genotyped using various generations of Illumina GWAS arrays. The genotypes were prephased using Eagle^73^ v2.3.5 and imputed using Beagle^74^ v4.1 with a reference panel of Finnish WGS data, the SISu v4.2 reference panel (*n* = 8,554). All the variants were processed on the human genome assembly GRCh38.

Clinical endpoints were defined based on medical records from multiple national health registries as previously described^34^. Laboratory measurements were obtained from the Kanta national laboratory database, which aggregates test results from healthcare providers across Finland. The Kanta lab dataset in FinnGen covers 2014–2023, with most data available from 2018 onwards. Of the 500,348 FinnGen participants, approximately 482,000 individuals had laboratory measurements in Kanta (average 482 tests per individual). Laboratory test results were harmonized to the Observational Medical Outcomes Partnership (OMOP) common data model^75^, with measurement units standardized and converted to OMOP conventions using harmonized values. The detailed pipeline of preprocessing and quality control of Kanta lab values are available at https://github.com/FINNGEN/kanta_lab_preprocessing.

### Sample collection

Peripheral blood mononuclear cell (PBMC) samples were obtained from FinnGen^34^ participants in collaboration with the Finnish Red Cross Blood Service as described elsewhere^36^. PBMCs were isolated from the buffy coat with Ficoll isogradient centrifugation. Following PBMC isolation, cells were resuspended in 10% DMSO in RPMI1640 medium (Gibco, 42401) supplemented with 5% inactivated human AB serum (Valley Biomedical Inc., HS1017HI), 2mM L-glutamine (Gibco, 25030-081), and 1% Pen-Strep (Gibco, 15140-12). The cell suspension was then aliquoted into cryovials (FluidX, 65-7641), placed in a CoolCell LX cell freezing container (Corning, 432138) and transferred into a –80 °C freezer. Cryovials were transferred into an automated gas phase liquid nitrogen freezer within 72 hours.

### PBMC nuclei isolation

Paired, single-nucleus gene expression and ATAC profiles were generated from PBMCs using the Chromium Next GEM Single Cell Multiome ATAC + Gene Expression assay (10x Genomics, PN-1000283). PBMCs were processed using the “Demonstrated Protocol Nuclei Isolation ATAC GEX Sequencing RevC” protocol (10x Genomics, Revision C, CG000365). Dead cells were removed via the EasySep Dead Cell Removal (Annexin V) protocol (Stem Cell Technologies, 17899). When applicable, remaining cells were used for FACS-based immunophenotyping.

For individual channels, cells were counted manually following Trypan staining (Thermo Fisher Scientific, 15250061). A total of one million live cells were carried forward to nuclei isolation. For multiplexed pooling, 16 samples were combined post-Dead Cell Removal at a total of 250,000 cells per sample. Cells were counted after AO/PI staining (Revvity Health Sciences Inc, CS2-0106-5ML) on the Cellometer Auto 2000 (Nexcelom Biosciences). A total of one million cells were carried forward to nuclei isolation. All nuclei were counted manually with a light microscope following DAPI staining (Sigma-Aldrich, D9542-1MG).

### Chromium Next GEM Single Cell Multiome ATAC + Gene Expression assay and sequencing

Following nuclei isolation, channels containing individual donors were loaded with 8,000–25,000 transposed nuclei. Multiplexed pools containing four, eight, or 16 donors were run over two, four, or eight channels, respectively, and were loaded with 20,000–40,000 nuclei. Samples underwent gene expression and ATAC library preparations were performed according to manufacturer instructions (10x Genomics). cDNA and ATAC libraries’ fragment sizes were assessed using the DNA High Sensitivity Bioanalyzer kit (5067-4626, Agilent). Gene Expression libraries’ fragment sizes were assessed using the High Sensitivity D1000 ScreenTape assay (Agilent, 5067-5585, 5067-5592). cDNA, ATAC and GEX libraries were quantified using the Qubit 1X dsDNA High Sensitivity assay kit (ThermoFisher Scientific,Q33231). Gene expression and ATAC libraries were sequenced on a variety of NovaSeq (SP, S2 and S4) and Novaseq (10B and 25B) flowcells (Illumina), in accordance with manufacturer instructions (10x Genomics, Illumina). Gene expression libraries were sequenced with the following read structure: Read 1: 28 cycles; Read 2: 90 cycles; i7 index read: 10 cycles; i5 index read: 10 cycles. ATAC libraries were sequenced with the following read structure: Read 1: 50 cycles; Read 2: 49 cycles; i7 index read: 8 cycles; i5 index read; 24 cycles.

### Sequencing data processing and quality control

We developed a computational pipeline to maximize the number of nuclei in our snRNA-Seq atlas and handle the mix of single donor and multiplexed channels (**Extended Data Fig. 1b**).

#### Single-nucleus RNA-sequencing (snRNA-Seq) pre-processing

Illumina BCL Convert v4.2.7 was used to demultiplex raw sequencing reads into FASTQ files for each channel. Cell Ranger^76^ count v7.0.1 was used with default settings to generate feature-barcode matrices. To eliminate ambient RNA and remove empty droplets, we applied CellBender^77^ remove-background v0.3 with default parameters.

#### Single-nucleus ATAC-sequencing (snATAC-Seq) pre-processing

Illumina BCL Convert v4.2.7 was used to demultiplex raw sequencing reads into FASTQ files for each channel. Cell Ranger ARC v2.0.2 was used to generate peak-barcode matrices and fragment files, with the maximum cell count limitation removed. All other Cell Ranger ARC parameters were set to default.

#### Genetic demultiplexing

For multiplexed samples, demuxlet^78^ was used to demultiplex nuclei based on imputed genotypes of 16 samples in each 16-plex pool. The pre-processed snRNA-seq and snATAC-seq BAM reads for each pool were merged and used jointly for demultiplexing, and only predicted singlets were retained for downstream analyses. CellBender-processed snRNA-Seq count matrices (filtered at false positive rate of 0.01) were then split by donor according to demuxlet results, creating donor-specific h5 files for each channel. Since pooled samples were run across multiple channels, this resulted in multiple h5 files per multiplexed donor.

#### snRNA-Seq quality control and filtering

Quality control (QC) and doublet prediction were performed using Cumulus^79^ v2.1.1 on each snRNA-Seq donor-channel h5 file. We applied the following exclusion criteria: nuclei with fewer than 400 unique molecular identifiers (UMIs), fewer than 200 detected genes, or greater than 20% of UMIs mapped to mitochondrial genes. Cumulus-predicted doublets were then filtered from each count matrix.

To efficiently annotate cell types at scale, we adopted Seurat’s reference-based label transfer approach. Each filtered count matrix was normalized using Seurat’s SCTransform, and cell type labels were transferred from the PBMC multimodal reference^37^, which contains cell type annotations at two levels of resolution: level 1 (L1) for broader cell types and level 2 (L2) for fine-grained cell types. Integration anchors between the query and reference datasets were identified using FindTransferAnchors (normalization.method = “SCT”, reference.reduction = “spca”, and dims = 1:50). Cell type labels were then transferred to the query dataset using TransferData, providing automated, scalable, and consistent annotations across all samples. To increase statistical power, we also defined a meta cell type called “PBMC” at the L1 level, which combines all nuclei across cell types for downstream analyses.

To remove low-quality nuclei, we applied an additional QC filter based on *MALAT1* expression^80^. *MALAT1* is a long non-coding RNA highly abundant in intact nuclei, and its expression level serves as a proxy for nuclear integrity. *MALAT1* thresholds were determined separately for each channel using an adaptive thresholding approach based on the bimodal distribution of SCTransform-normalized MALAT1 counts. Nuclei with MALAT1 expression below the channel-specific empirically determined threshold were excluded prior to integration.

#### snRNA-seq sample integration

Demultiplexed, filtered snRNA-Seq data from all demultiplexed sample-channels and single-donor channels were combined into a single atlas. To efficiently handle millions of nuclei, the integrated expression matrix was created using Scanpy^81^ v1.10.1 and anndata^82^ v0.10.7, combining CellBender-processed count matrices with Seurat-based cell type annotations.

For dimensionality reduction and visualization, the resulting expression matrix was processed using Scanpy. Library size normalization was performed with a target sum of 10,000 counts per nucleus, followed by natural log transformation. Highly variable genes were identified with default parameters, and the data was subset to these genes and scaled (max_value = 10). Principal component analysis (PCA) was performed (svd_solver = “arpack”), followed by neighborhood graph construction and UMAP embedding with default parameters.

#### snATAC-seq quality control and filtering

Per-channel snATAC-seq peak-barcode matrices were first subset to barcodes retained in the snRNA-seq atlas, leveraging established empty droplet and doublet prediction approaches to ensure all snATAC-seq nuclei had paired snRNA-seq data. For peak calling, we computed standard QC metrics for each sample using per-sample peaks called by Cell Ranger ARC and Signac^83^ v1.14.0, retaining nuclei with the following criteria: 2,000 < the number of fragments in peaks < 100,000, the ratio of reads in the ENCODE’s genomic blacklist regions < 0.01, nucleosome signal < 2, and TSS enrichment score > 4. We then split fragments by L2 cell types using Signac’s SplitFragments.

#### snATAC-seq consensus peak calling

For each L2 cell type, we called pseudobulk cell-type-specific peaks using MACS2 v2.2.9.1 (ref. ^38^). We first merged post-QC per-sample cell-type-specific fragment files across samples to assemble pseudobulk cell-type-specific fragments. To leverage paired-end reads from snATAC-seq, we converted these pseudobulk fragments into the tagAlign format following the Activity-by-Contact (ABC) model pipeline^84^. We then called pseudobulk cell-type-specific narrow peaks and peak summits using MACS2 with the parameters as implemented in the ABC model pipeline: peak *P*-value threshold of 0.1, shift of the 5’ end of all reads toward 3’ by 75 bp, and 150 bp extension of them in the 5’ direction, consistent with the ENCODE ATAC-seq pipeline. For each cell type, we retained the top 150,000 narrow peaks ranked by overlapping read counts, defined 500 bp elements centered on the peak summits, merged overlapping elements, and removed any elements overlapping with the ENCODE’s blacklist regions. To create a consensus set of peaks across cell types, we applied ArchR’s iterative overlap peak merging approach, which iteratively retains non-overlapping cell-type-specific peaks sorted by read counts^85,86^.

#### snATAC-seq sample integration

To remove reads showing reference allele mapping bias, we applied WASP^87^ filtering as implemented in the SALSA workflow^88^ to BAM files processed by Cell Ranger ARC, generating WASP-corrected fragment files per sample. We used per-sample imputed genotypes for WASP, retaining only heterozygous single-nucleotide variants (SNVs) with minor allele count (MAC) > 20 among the single-cell donors.

We then created individual-level peak-barcode count matrices based on the consensus peaks using Signac to quantify chromatin accessibility by counting fragments overlapping each peak per nucleus. We recomputed the standard QC metrics for each sample using the WASP-corrected count matrices and applied the same QC criteria as described above. To handle millions of nuclei, we merged the filtered count matrices across samples using Scanpy and anndata, then annotated cell types transferred from the snRNA-seq atlas. For dimensionality reduction and visualization, we used SnapATAC2 v2.8.0 (ref. ^89^). We first selected the top 50,000 highly variable peaks across the dataset and computed spectral embeddings using cosine distance without feature weighting. We then generated UMAP coordinates from the spectral embeddings for visualization.

### Peak–gene link analysis

To identify enhancer–gene regulatory links from paired chromatin accessibility and gene expression data, we employed a hurdle model framework that accounts for zero-inflation and overdispersion characteristics of single-cell expression data, as originally implemented in Open4Gene^32^, which has been successfully applied to identify peak–gene links with well-calibrated performance. We modeled the association between chromatin accessibility and gene expression using a two-part hurdle model that independently fits 1) a binomial zero-inflation model testing whether peak accessibility affects the probability of nonzero expression, and 2) a negative binomial count model testing whether peak accessibility affects expression magnitude among expressing nuclei. This approach explicitly accounts for technical and biological sparsity while modeling the count-based nature of expression measurements with overdispersion, making it well-suited for single-cell data.

For each cell type, we systematically tested associations between gene expression and chromatin accessibility of consensus peaks in *cis*. We generated candidate peak–gene pairs based on genomic proximity, pairing each gene with all consensus peaks within 1 Mb of its TSS. To focus on biologically relevant associations and improve computational efficiency, we filtered peak–gene pairs using two criteria: 1) genes and peaks with FDR < 0.05 for *cis*-eQTL or *cis*-caQTL effects (*i.e.,* eGenes and caPeaks); and 2) genes and peaks with nonzero expression or accessibility proportion of ≥ 1% in cells from the tested cell type, ensuring sufficient statistical power.

For each filtered peak–gene pair, we fitted the hurdle model with cell-level covariates including log-transformed total RNA and ATAC counts, and the percentage of mitochondrial counts per cell. To control for highly expressed genes or peaks that could dominate normalization, genes or peaks exceeding 5% of total counts in any cell were excluded when computing size factors. To efficiently handle millions of nuclei, we implemented an optimized Rcpp-based hurdle model called fasthurdle (https://github.com/mkanai/fasthurdle), which is ∼13× faster than the original R implementation (pscl) for use of binomial zero-inflation and negative binomial count models^32^. Given the size of our dataset, we were unable to apply alternative methods such as SCENT^30^, which requires substantial computational burden for adaptive permutations.

We obtained *P* values from both of the zero-inflation and count models. To control FDR while accounting for the two-stage nature of the hurdle model, we applied a Jiang-Doerge procedure^90^ adapted for paired hypothesis testing. In the first stage, we identified peak–gene pairs with relatively lenient filtering using Benjamini-Hochberg FDR correction on zero-inflation *P* values (*α*_1_= 0.1). In the second stage, we estimated the proportion of true nulls among selected pairs and applied Benjamini-Hochberg correction to count model *P* values with an adjusted threshold (*α*_2_ = 0.05) that accounts for the initial selection. Peak–gene links were considered significant for a given cell type if they passed both stages of this procedure.

### Co-accessibility analysis

To examine coordinated chromatin accessibility patterns at regulatory hotspots, we performed co-accessibility analysis at the 19p13.11 locus (containing *IFI30*, *JUND*, *MAST3*, and *IL12RB1*) and the 8p21.3 locus (containing TRAIL receptors, *TNFRSF10A–D*). For each locus, we extracted pseudobulk chromatin accessibility profiles across all consensus peaks in the region and computed pairwise Pearson correlations. For visualization, median correlation coefficients were computed in a 256 × 256 grid and displayed as co-accessibility maps. To quantify the density of peak–gene links, we also computed kernel density estimates of linked peak positions for each linked gene using 512 bins.

### Expression quantitative trait locus (eQTL) mapping

#### Pseudobulk analysis

Pseudobulk expression per gene was computed by taking the mean of log2-transformed, count-per-ten-thousand (CP10K) normalized expression matrices for each cell type. We used the mean expression because this metric is most comparable with the SAIGE-QTL framework^16^. For a given cell type, we excluded samples with less than 10 cells and genes expressed in less than 50% of the samples. We then inverse-rank normalized pseudobulk mean expression to use in pseudobulk eQTL analysis.

For each cell type, the optimal number of PEER factors^91^ was determined by testing 0–60 PEER factors and selecting the value that maximized the number of *cis*-eGenes at false discovery rate (FDR) < 0.05, following recommendations for single-cell eQTL studies^92^. PEER factors were computed using peerR v1.3 with covariates including age at sample collection, sex, and top four genetic PCs.

Pseudobulk eQTL mapping was conducted using tensorQTL^41^ v1.0.10 for each cell type with covariates including age, sex, top four genetic PCs, and cell-type specific PEER factors as computed above (**Supplementary Table 33**). For *cis*-eQTL analysis, we tested all imputed variants within 1 Mb of the transcription start site (TSS) with MAC > 20 among the single-cell donors. To compute gene-level *P* values, we used the ACAT-V test^93^ to combine variant-level nominal *P* values using a Cauchy combination, instead of tensorQTL’s default beta-approximated *P* values^94^, which enables a direct comparison with SAIGE-QTL^16^. We then computed FDR using Storey’s method^95^ and defined *cis*-eGenes with FDR < 0.05. For *trans*-eQTL analysis, we tested all imputed variants with MAF > 5% that are located outside of the cis window (*i.e.*, > 5 Mb away or on different chromosomes). We defined *trans*-eGenes as those having at least one genome-wide significance variant (*P* < 5.0 × 10^-8^).

#### SAIGE-QTL

Single-cell *cis*-eQTL analysis was conducted using SAIGE-QTL^16^ v0.3.0 for each cell type. We ran step 1 of SAIGE-QTL using the following parameters and covariates: tol = 0.0001, maxiterPCG = 50000, isCovariateOffset = FALSE, and useGRMtoFitNULL = FALSE. Sample-level covariates include age at sample collection, sex, the top four genetic PCs, and cell-type specific PEER factors as computed in the pseudobulk analysis (**Supplementary Table 33**). Cell-level covariates included log-transformed total RNA counts, and the percentage of mitochondrial counts per cell. To control for highly expressed genes that could dominate normalization, genes exceeding 5% of total counts in any cell were excluded when computing size factors. For a given cell type, we ran SAIGE-QTL for genes that we retain in the pseudobulk analysis (*i.e.*, genes expressed in more than 50% of the samples at the pseudobulk level). We next ran step 2 of SAIGE-QTL using imputed genotypes with MAC > 20. Gene-level *P* values and FDR were calculated from variant-level *P* values using the ACAT-V test^93^ and Storey’s method^95^, respectively, as described above. Given the scale of our dataset, *trans*-eQTL was not conducted for this project.

### Chromatin accessibility quantitative trait locus (caQTL) mapping

Pseudobulk chromatin accessibility per peak was computed by taking the sum of log2-transformed, count-per-ten-thousand (CP10K) normalized accessibility matrices for each cell type. We used the sum of accessibility counts to address the inherent sparsity of snATAC-seq data, where most nuclei show zero counts at a given peak. We then applied the same filtering criteria for peaks and conducted pseudobulk caQTL analyses using tensorQTL^41^, following the same procedure as the pseudobulk eQTL analysis with the following differences: 1) instead of PEER factors, we computed the optimal number of top PCs from covariate-residualized (age, sex, top four genetic PCs) pseudobulk chromatin accessibility matrices by testing 0–100 PCs that maximized the number of discovered caPeaks, because the application of PEER factors for chromatin accessibility peaks has not been fully explored; 2) for *cis*-caQTL analysis, we tested variants within 100 kb of the peak center.

### Protein quantitative trait locus (pQTL) mapping

Protein abundance was quantified using the Olink Explore 3072 platform across three batches, comprising 2,190 total samples from FinnGen. QC removed protein abundance PC outliers (mean ± 5SD), median and interquartile range (IQR) outliers, duplicated samples, sex mismatches, and inter-batch duplicates, retaining 1,990 samples and 2,925 proteins. After further removal of genetically related individuals, 1,732 unrelated samples remained for pQTL analysis, of which 680 samples were collected through the Finnish Red Cross Blood Service (as described elsewhere^36^) and have QC-passing single-nucleus multiome measurements.

Protein measurements were log-transformed and residualized for age at sample collection, sex, batch, plate, row, and five genetic PCs, followed by inverse-rank normalization. Genome-wide pQTL mapping was conducted using PLINK2 v2.00a3.3LM^96^ with all imputed variants with MAC > 8. For *cis*-pQTL analysis, we tested variants within 1 Mb of the gene’s TSS.

### Genome-wide association study (GWAS) of complex traits

Of 2,857 complex trait GWAS available in FinnGen R12 (2,466 binary endpoints, three anthropometric traits, 383 laboratory measurements, and five lab-derived traits), we analyzed 439 complex traits with sufficient statistical power (*Z* > 2 for *h*^2^g estimate by LDSC^97^ and number of genome-wide significant loci > 10; **Supplementary Table 23**). GWAS was performed using REGENIE^98^ v3.3 with a two-step procedure via FinnGen’s REGENIE pipeline (https://github.com/FINNGEN/regenie-pipelines). In step 1, genetic relatedness was computed using 188,153 well-imputed, LD-independent common variants (INFO > 0.95 across all batches, >97% non-missing genotypes, MAF > 1%, and *r*^2^ < 0.2 within 1.5 Mb window) to compute leave-one-chromosome-out (LOCO) predictions for each phenotype. In step 2, association tests were conducted for variants with MAC ≥ 5 among cases and controls using approximate Firth regression for variants with initial *P* < 0.01. Covariates included sex, age at the end of follow-up or death, genotyping batch, and the top 10 genotype PCs. Summary statistics were then filtered by INFO > 0.6.

For laboratory measurements, we conducted GWAS only for those with at least 1,000 individuals having one or more measurements. Sample-wise median values were calculated and inverse-rank normalized for quantitative laboratory measurements (*n* = 347), while binary laboratory traits (*n* = 36) were defined as ever versus never having measurements above specific thresholds or test outcomes classified as abnormal or high. For quantitative laboratory measurements, we applied an additional filter of MAC > 30 to control for inflation of test statistics observed among very low allele count variants.

### Statistical fine-mapping

#### eQTLs and caQTLs

For each pair of eGenes or caPeaks (FDR < 0.05) and cell types, we conducted statistical fine-mapping using SuSiE^99^ v0.14.2 with *cis*-eQTL or *cis*-caQTL summary statistics and covariate-adjusted in-sample dosage LD matrices. We used *cis*-regions of eQTLs or caQTLs (1 Mb around TSS or 100 kb around the peak center) as defined in the molQTL analyses. For *cis*-eQTL, we used SAIGE-QTL summary statistics to maximize power. Allowing up to 10 causal variants per eGene or caPeak, we derived up to 10 independent 95% credible sets (CS) and calculated posterior inclusion probabilities (PIP) of each variant using the default uniform prior probability of causality.

Fine-mapping methods typically assume linear models without covariates^99,100^, whereas GWAS and molQTL analyses include covariates. Using the Frisch-Waugh-Lovell (FWL) theorem^101^, covariate-adjusted summary statistics can be paired with covariate-adjusted in-sample LD for fine-mapping^7,68,102^. This adjustment is important when genotypes are correlated with covariates (*e.g.*, genetic PCs capturing population stratification) and more critical for molQTL analyses where PEER factors or expression PCs capture both technical confounders and genetically regulated global expression patterns^91^.

We therefore adjusted in-sample dosage LD matrices using the same sample-level covariates as in the molQTL analyses (age at sample collection, sex, top four genotype PCs, and cell-type-specific PEER factors for eQTLs or accessibility PCs for caQTLs). We developed ldcov (https://github.com/mkanai/ldcov) to efficiently compute covariate-adjusted LD using the FWL theorem. After L2 normalization of genotypes, we applied QR decomposition-based FWL projection, which is numerically more stable than direct pseudoinverse computation when covariates are collinear. For computational efficiency, we pre-computed the orthogonal projection matrix Q from QR decomposition once per cell type and reused it across loci, since the projection depends only on covariates, not genotypes.

#### pQTLs and complex trait GWASs

Because pQTLs and complex trait GWASs were originally conducted genome-wide, we used FinnGen’s standard fine-mapping pipeline (https://github.com/FINNGEN/finemapping-pipeline) as described previously^68^. Briefly, we defined fine-mapping regions based on a 3 Mb window around each lead variant and merged overlapping regions. For each region, we conducted statistical fine-mapping using SuSiE^99^ v0.11.92 with summary statistics and in-sample dosage LD matrices without covariate-adjustment. For *cis*-pQTL analysis, we only retained fine-mapped 95% CSs within 1 Mb of each gene’s TSS.

### Colocalization

We performed colocalization within and across molQTLs using a SuSiE^99^ fine-mapping-based colocalization approach as implemented in the coloc framework^103^, using FinnGen’s colocalization pipeline (https://github.com/FINNGEN/coloc.susie.direct). Each CS was defined by a combination of molQTL type, molecular feature (gene or peak), and cell type. Within each molQTL type, we colocalized CSs for the same gene or peak across cell types to identify shared genetic regulation. Across molQTL types, we colocalized CSs among overlapping eQTLs, caQTLs, and pQTLs. For each pair of overlapping CSs, we computed the posterior probability of colocalization (PP.H4), representing the probability that both traits share the same causal variant in a given region. We applied the following filtering thresholds for colocalization: PP.H4 > 0.5, CS log_10_ Bayes factor > 0.9, minimum probability mass in both CSs in the overlapping region > 0.5, and number of overlapping variants in CSs > 0.

To systematically merge pairwise collocalized CSs across cell types and molQTLs, we implemented a two-step graph-based clustering approach: 1) partitioning CSs into connected components based on pairwise colocalization relationships, and 2) within each component, clustering CSs by shared variants to identify maximal subsets sharing variants. Singleton CSs were retained as individual merged CSs.

We further performed colocalization analysis between molQTLs and the 439 GWAS traits from FinnGen R12 using the same fine-mapping-based approach described above.

### Comprehensive Analysis Suite for Cell-type specificity and Accessibility-Driven QTL Effects (CASCADE)

We developed CASCADE (Comprehensive Analysis Suite for Cell-type specificity and Accessibility-Driven QTL Effects) to systematically characterize cell type specificity and regulatory mechanisms of molecular QTLs (https://github.com/mkanai/cascade). CASCADE implements a hierarchical framework that operates at two levels: 1) gene-or peak-level analysis to classify eGenes and caPeaks by their cell type specificity patterns, incorporating power-aware assessment through multivariate adaptive shrinkage (mash)^43^; and 2) variant-level analysis to classify individual fine-mapped variants by their regulatory mechanisms (chromatin-to-expression cascades) and cell type specificity.

#### Multivariate adaptive shrinkage (mash) analysis

To distinguish true cell type specificity from limited statistical power, we applied mash^43^ to jointly model effect sizes across cell types. Mash leverages patterns of effect sharing across conditions to improve power for detecting effects and estimate local false sign rates (LFSR), which quantify confidence in the sign and magnitude of effects.

For each molecular feature (gene or peak), we constructed matrices of effect sizes and standard errors across all cell types and variants. We selected three subsets of variants: 1) *strong* variants with fine-mapped CSs (PIP > 0.1) or, as an alternative when no CSs available, the top five variants with the strongest association (*Z* > 4.89) across any cell type; 2) *null* variants comprising five randomly selected LD-independent variants with weak associations (*Z* < 2.0) across all cell types; and 3) *random* variants comprising five randomly selected LD-independent variants. Missing effect estimates in any cell types were set to zero with large standard errors (= 1000).

We estimated the residual correlation structure among cell types using null variants. We then computed data-driven covariance matrices from strong variants using the mash’s standard procedure: FLASH (with non-negative factor loadings) and PCA (five components), followed by extreme deconvolution to refine estimates, combined with canonical covariance matrices. We fit the mash model in two stages: first estimating mixture weights using random variants with data-driven and canonical covariance matrices, then computing posterior summaries for strong variants with fixed weights. From the fitted model, we extracted LFSR for each gene-or peak-variant pair in each cell type, representing the posterior probability that the effect has the opposite sign from the estimated effect.

#### Cell-type hierarchy and annotation levels

Cell type annotations were performed at two hierarchical levels using Seurat’s reference-based label transfer as described above. L1 annotations define 9 broad cell types: PBMC, monocytes (Mono), dendritic cells (DC), natural killer cells (NK), B cells, CD4+ T cells, CD8+ T cells, other T cell subtypes, and other cell types. L2 annotations provide finer-grained resolution. For CASCADE analyses, we leveraged QTL mapping performed at both L1 and L2 cell types. When analyzing L2 cell types, we mapped them to their corresponding L1 parent types to maintain hierarchical consistency in specificity classification as follows:

- **Mono**: CD14^+^ and CD16^+^ monocytes
- **DC**: conventional type 1 and 2 DCs (cDC1 and cDC2), and plasmacytoid DCs (pDC)
- **B**: Intermediate, memory, naive B cells, and plasmablasts
- **CD4^+^ T**: CD4^+^ cytotoxic T lymphocytes (CD4^+^ CTL), CD4^+^ naive T cells (CD4^+^ Naive), CD4^+^ central memory T cells (CD4^+^ TCM), CD4^+^ effector memory T cells (CD4^+^ TEM), and regulatory T cells (Treg)
- **CD8^+^ T**: CD8^+^ naive T cells (CD8^+^ Naive), and CD8^+^ effector memory T cells (CD8^+^ TEM)
- **NK**: NK, CD56^bright^ NK cells (NK CD56^bright^), and proliferating NK cells (NK Proliferating)
- **Other T**: mucosal-associated invariant T cells (MAIT), double-negative T cells (dnT), and gamma-delta T cells (gdT)
- **Other**: innate lymphoid cells (ILC), hematopoietic stem/progenitor cells (HSPC), and platelets

This hierarchical structure allows CASCADE to classify cell type specificity based on L1 lineages (myeloid vs. lymphoid) while incorporating information from finer L2 subtypes when available. During specificity categorization, L2 cell types are aggregated to their L1 parents to determine lineage-level and cross-lineage sharing patterns.

#### Cell-type specificity of eGenes and caPeaks

Using mash-derived LFSR values, we classified eGenes and caPeaks based on their cell type specificity. We first identified features with significant effects (FDR < 0.05) across cell types, then used LFSR values to distinguish true specificity from limited statistical power. Based on patterns of FDR significance and LFSR thresholds (LFSR < 0.05 for likely shared effects, LFSR ≥ 0.5 for likely specific effects), we classified eGenes and caPeaks into five hierarchical categories that account for immune cell lineage structure:

1. **Cross-lineage shared**: Significant (FDR < 0.05) in both myeloid (Mono, DC) and lymphoid (NK, B, CD4^+^ T, CD8^+^ T, other T cells) lineages
2. **Likely shared but underpowered**: Significant in one lineage (or only PBMC) but with low LFSR (< 0.05) across other lineages, indicating likely shared effects that lack statistical power
3. **Lineage-specific**: Significant in either myeloid or lymphoid lineage only, with high LFSR (≥ 0.5) in the other lineage
4. **T-cell-specific**: Significant only in T cell subtypes (CD4^+^ T, CD8^+^ T, and other T cells)
5. **Single cell-type**: Significant in one cell type

For eGenes and caPeaks with multiple fine-mapped CSs, we analyzed whether effects across cell types arose from the same or distinct causal variants using the merged CS clusters described above. For each CS cluster, we identified the top variant with the maximum *χ*^2^ value and assessed effect size heterogeneity across cell types using Cochran’s Q heterogeneity test (*P* < 5.0 × 10^-8^). We classified variant sharing patterns into four heterogeneity categories with the following priority order:

1. **Shared with opposite effects**: At least one CS cluster shows same causal variant(s) with effects in opposite directions across cell types (Cochran’s *Q p*-value < 5.0 × 10^-8^)
2. **Shared with heterogeneous effects**: At least one CS cluster shows same causal variant(s) with heterogeneous effect sizes in the same direction (Cochran’s *Q p*-value < 5.0 × 10^-8^)
3. **Shared with consistent effects**: All CS clusters show same causal variant(s) with homogeneous effect sizes (Cochran’s *Q p*-value ≥ 5.0 × 10^-8^)
4. **Distinct variants**: Multiple CS clusters affecting different cell types with no shared CSs

#### QTL mechanisms of variants

We next classified individual fine-mapped molQTL variants (PIP > 0.5) based on their regulatory mechanisms to identify complete chromatin-to-expression cascades. For each variant in each cell type, we evaluated four key features:

1. **Chromatin accessibility peak overlap**: Whether the variant overlaps a consensus peak accessible in that cell type
2. **caQTL status**: Whether the variant shows caQTL effects in that cell type for any peak, including both overlapping and non-overlapping peaks
3. **Peak–gene links**: Whether peaks affected by the variant (either overlapping or caQTL peaks) have peak–gene links in that cell type
4. **eQTL status**: Whether the variant shows eQTL effects in that cell type for any gene, and whether the affected gene is linked to a variant-associated peak

Based on combinations of these four features, we classified variants into 25 mutually exclusive patterns (**Extended Data Fig. 6a**), which we then collapsed into six major mechanistic categories:

1. **Full Cascade**: Variant shows caQTL effects, the affected peak has a peak–gene link, and the variant shows eQTL effects for the linked gene, representing a complete regulatory cascade from chromatin accessibility to gene expression
2. **Positional Cascade**: Variant overlaps a peak with a peak–gene link and shows eQTL effects for the linked gene, but does not show detectable caQTL effects
3. **caQTL + eQTL (No Link)**: Variant shows both caQTL and eQTL effects, but the affected gene lacks a peak–gene link to the caQTL peak, or the eQTL gene differs from the linked gene
4. **Only caQTL (With Link)**: Variant shows caQTL effects and the affected peak has peak–gene links, but no eQTL is detected for the linked gene
5. **Only caQTL (No Link)**: Variant shows caQTL effects but the affected peak lacks peak–gene links
6. **Only eQTL**: Variant shows eQTL effects but no detectable caQTL effects, regardless of peak overlap or links

We defined a seventh category, “No molQTL”, for completeness, representing variants showing neither caQTL nor eQTL effects in any cell type analyzed. However, this category was not assigned in our CASCADE analysis because we only classified fine-mapped molQTL variants (PIP > 0.5 in either eQTL or caQTL). We first assigned QTL mechanism patterns in each cell type, then aggregated them across cell types by taking the highest-priority mechanism for each variant.

#### Cell-type specificity of variants

We assessed variant-level cell type specificity using the same hierarchical classification framework applied to eGenes and caPeaks. For each variant, we first determined which cell types exhibited the variant’s highest-priority QTL mechanism (lowest pattern number). We then applied the five-category classification based on the distribution of highest-priority effects across cell types. This approach ensures that cell type specificity assessment focuses on the most complete regulatory mechanisms rather than partial or indirect effects. Because the mash analysis focused on sharing of the top effects, the “Likely shared but underpowered” category was only assigned to variants with the highest-priority QTL mechanism in PBMC alone.

#### Enrichment of GWAS colocalizing variants

To assess the disease relevance of different regulatory mechanisms, we examined GWAS colocalization rates across CASCADE mechanistic categories. For each fine-mapped molQTL variant, we determined whether it colocalized with any of the 439 FinnGen GWAS traits as described above, then calculated colocalization rates as the proportion of variants in each mechanistic category showing GWAS colocalization.

As a negative control, we analyzed LD-pruned variants (*r*^2^ < 0.2) with no detected molQTL effects (maximum PIP < 0.01 in both eQTL and caQTL), sampled from the same genomic regions to control for regional confounding. For comparison with pQTLs we incorporated *cis*-pQTL results (PIP > 0.5) and calculated colocalization rates for: 1) any pQTL variants regardless of CASCADE status, and 2) pQTL-only variants without detectable caQTL or eQTL effects.

### Functional enrichment

We performed functional enrichment analysis for chromatin accessibility peaks with significant peak–gene links and for fine-mapped molQTL variants by annotating peaks or variants with functional annotations. For both analyses, we estimated functional enrichment as relative risk (i.e., a ratio of proportions) by comparing the features with the annotation in the test group vs. the background group^68^, and calculated 95% confidence intervals (CIs) using bootstrapping with 1,000 replicates.

#### Chromatin accessibility peaks with significant peak–gene links

We used ENCODE4 candidate *cis*-regulatory elements (cCREs)^40^ to annotate overlapping peaks with the following classifications: promoter-like signatures (PLS), proximal enhancer-like signatures (pELS), distal enhancer-like signatures (dELS), chromatin accessible with CTCF (CA-CTCF), chromatin accessible with H3K4me3 (CA-H3K4me3), chromatin accessible with TF (CA-TF), chromatin accessible only (CA), and transcription factor only (TF). When a peak spanned across multiple cCRE classifications, we assigned a “Mixed” annotation. We compared peaks with at least one significant peak–gene link across cell types (test group) to those without links (background group). Relative risk = (proportion of linked peaks overlapping the annotation) / (proportion of non-linked peaks overlapping the annotation).

#### Fine-mapped molQTL variants

We defined eight distinct functional categories in sequential order: pLoF (predicted loss-of-function), missense, synonymous, intron, 5’ untranslated region (UTR), 3’ UTR, cCRE, and non-cCRE. Variant-based categories (pLoF, missense, synonymous, intron, and 5’/3’ UTR variants) were defined based on the most severe consequence for each variant on a canonical transcript, as predicted by the Ensembl Variant Effect Predictor (VEP)^104^ v105 (using GRCh38 and GENCODE v39). The pLoF category comprised stop-gained, splice site disrupting, and frameshift variants predicted as high-confidence by LOFTEE^62^. The missense category included missense-like variants such as low-confidence LoF variants. Region-based categories (cCRE and non-cCRE) were then defined based on overlap with any of the ENCODE4 classifications above. We also annotated each variant by its cCRE classification (independent of VEP annotations), and further stratified coding variants (pLoF, missense, and synonymous) by cCRE overlap. To investigate enrichment of peaks significantly associated with gene expression, we additionally stratified variants by overlap with linked peaks.

For each variant, we computed the maximum PIP across *cis*-eQTLs or *cis*-caQTLs, then compared fine-mapped variants (PIP > 0.9, test group) to background variants (PIP ≤ 0.01). Relative risk = (proportion of variants with PIP > 0.9 in the annotation) / (proportion of variants with PIP ≤ 0.01 in the annotation).

### Mediated Expression Score Regression (MESC)

To quantify the proportion of disease heritability mediated by molecular traits, we applied mediated expression score regression (MESC)^24^. MESC uses individual-level genotypes and molecular phenotypes to construct expression scores that capture the genetic component of molecular trait variation, which are then regressed against GWAS summary statistics to estimate QTL-mediated heritability (*h*^2^_med_). To evaluate the joint contribution of multiple molecular layers, we performed MESC analyses on seven molQTL configurations: caQTLs alone, eQTLs alone, pQTLs alone, and all pairwise and three-way combinations (*i.e.*, caQTL + eQTL, caQTL + pQTL, eQTL + pQTL, caQTL + eQTL + pQTL), as described previously^25^.

For each cell type and molQTL, we computed expression scores separately using molecular phenotype matrices, matching in-sample genotypes, and the SISu v4.2 reference panel (*n* = 8,554) as the LD reference. We used the same phenotype matrices and covariates as in the pseudobulk eQTL, caQTL, or bulk plasma pQTL analyses. For the combinations of molQTLs, we restricted analysis to overlapping samples. We applied MESC using the 439 complex trait GWAS summary statistics from FinnGen (R12), jointly with the modified baselineLD v2.0 reference provided by MESC. This reference includes 72 functional annotations, excluding four QTL MaxCPP annotations^105^ from the original v2.0 reference^106^. We computed *h*^2^_med_ / *h*^2^_g_, representing the proportion of disease heritability mediated by each molQTL or combination. To investigate enrichment patterns across disease categories (immune, cancer, and cardiometabolic), we performed random-effects meta-analysis (REML) of *h*^2^_med_ / *h*^2^_g_ across traits within each category.

### Stratified LD score regression (S-LDSC)

We applied stratified LD score regression (S-LDSC)^61^ to quantify disease heritability and its enrichment by chromatin accessibility annotations, regardless of QTL-mediated effects. We computed LD scores for three chromatin accessibility annotation sets per cell type: 1) all chromatin accessibility peaks, 2) peaks with peak–gene links (CA-Link+), and 3) peaks without links (CA-Link–). LD scores were calculated using the 1000 Genomes European ancestry reference panel (GRCh38) restricted to HapMap3 SNPs, following the standard S-LDSC pipeline.

For each chromatin accessibility annotation, S-LDSC was performed using the 439 complex trait GWAS summary statistics from FinnGen (R12), jointly with the baselineLD v2.2 reference that includes 97 functional annotations^107^. We computed the proportion of heritability (*h*^2^_g_) and heritability enrichment (*h*^2^_g_ / the proportion of SNPs in the annotation). To investigate enrichment patterns across disease categories (immune, cancer, and cardiometabolic), we performed random-effects meta-analysis (REML) across traits within each category, separately for proportion of heritability and enrichment statistics.

### Constraint analysis

To characterize the regulatory architecture across the constraint spectrum, we used fine-mapped molQTL variants (PIP > 0.5) and loss-of-function observed/expected upper bound fraction (LOEUF) deciles from gnomAD^62,63^ v4.1. LOEUF deciles were assigned to 15,976 protein-coding genes expressed in PBMCs, and we computed Spearman’s correlations between LOEUF deciles and the corresponding effect sizes for each gene.

For eQTL and caQTL analyses, we selected the fine-mapped variant with the maximum absolute effect size per gene or linked peak, respectively. For peak–gene link analyses, we used significant peak–gene links and quantified both the number of linked peaks per gene and the maximum absolute effect size of linked peaks per gene from the negative binomial count component of the hurdle model. To approximate the combined regulatory effect of genetic variants on gene expression through chromatin accessibility (*β*_combined_), we multiplied caQTL effect sizes (*β*_caQTL_) by peak–gene link effect sizes (*β*_link_) for each caQTL–peak–gene combination, taking the maximum absolute *β*_combined_ = *β*_caQTL_ × *β*_link_ across all peaks linked to the gene.

We note that these effect sizes are derived from different statistical models operating on different molecular features and therefore have different units: *β*_eQTL_ represents change in log gene expression counts per allele (SAIGE-QTL^16^ Poisson mixed model); *β*_caQTL_ represents change in pseudobulk chromatin accessibility in standard deviation (s.d.) units per allele (tensorQTL^41^ Gaussian model); *β*_link_ represents change in log gene expression counts per unit increase in chromatin accessibility counts (negative binomial count component of the hurdle model^32^); and *β*_combined_ = *β*_caQTL_ × *β*_link_ serves as a relative measure of combined regulatory effect, with units not directly comparable to *β*_eQTL_. Although absolute effect sizes cannot be directly compared across these models, the constraint-dependent gradient (*i.e.*, Spearman’s correlation between LOEUF deciles and effect sizes) remains valid for comparing *β*_eQTL_ and *β*_combined_.

To control for the relationship between gene expression levels and constraint, we additionally analyzed expression-matched highly-expressed genes by: 1) filtering for genes in the top 20% of mean expression levels across PBMC pseudobulk samples, 2) stratifying genes into 10 × 10 bins based on LOEUF decile and mean expression decile, and 3) randomly sampling genes from each bin to match the minimum number of genes across all bins within each expression decile. Mean expression and expression variance were calculated from pseudobulk CP10K count matrices for PBMCs. Expression variance residuals were computed by regressing log-transformed variance on log-transformed mean expression using LOESS regression (span = 0.1) as described previously^64^.

To assess peak constraint, we used genomic non-coding constraint of haploinsufficient variation (Gnocchi) scores from gnomAD^63^ v3. For each gene with linked peaks, we identified three representative peaks: 1) the most constrained peak (using the same approach as the Gnocchi manuscript^63^), 2) the strongest effect peak, and 3) a randomly selected peak among linked peaks per gene. We then examined how the Gnocchi scores of these peaks varied across LOEUF deciles. For stratified analyses by both peak and gene constraint, we categorized peaks as “constrained” if Gnocchi > 4 or “unconstrained” if –4 < Gnocchi < 4.

For GWAS colocalization enrichment analyses across LOEUF deciles, we calculated the proportion of genes in each LOEUF decile that showed colocalization (PP.H4 > 0.5) between eQTLs or caQTLs at linked peaks and at least one of the 439 GWAS traits from FinnGen R12.

## Extended Data Figures

**Extended Data Fig. 1.**
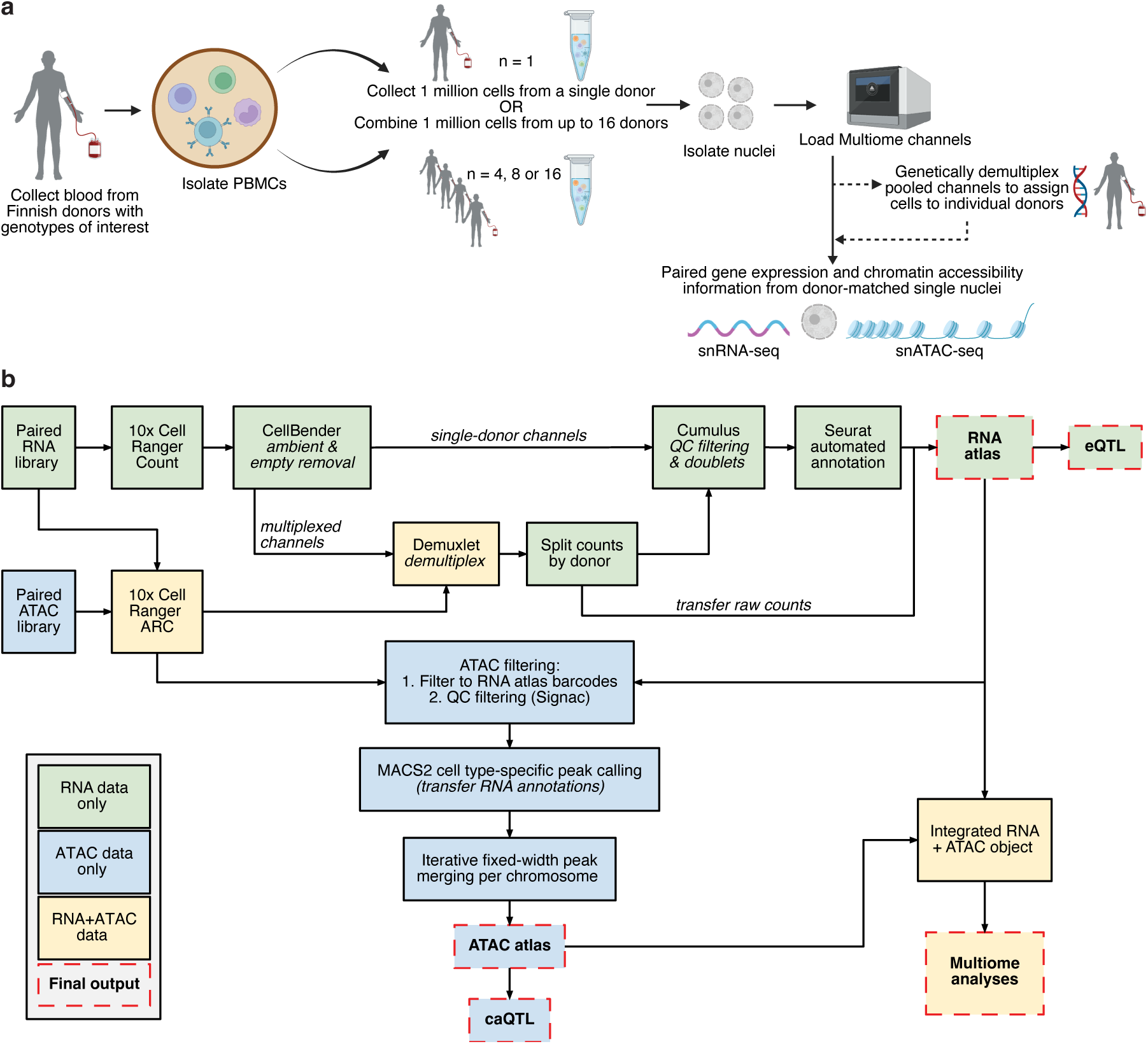
Schematic overview of experimental and computational pipelines a, Sample processing workflow. Peripheral blood mononuclear cells (PBMCs) from 1,171 donors were processed to generate paired single-nucleus RNA sequencing (snRNA-seq) and single nucleus assay for transposase-accessible chromatin sequencing (snATAC-seq) data. **b,** Multiome analysis pipeline. Data was used to generate snRNA-seq and snATAC-seq atlases, and an integrated object for multiome analyses. QC, quality control. eQTL, expression quantitative trait locus.

**Extended Data Fig. 2.**
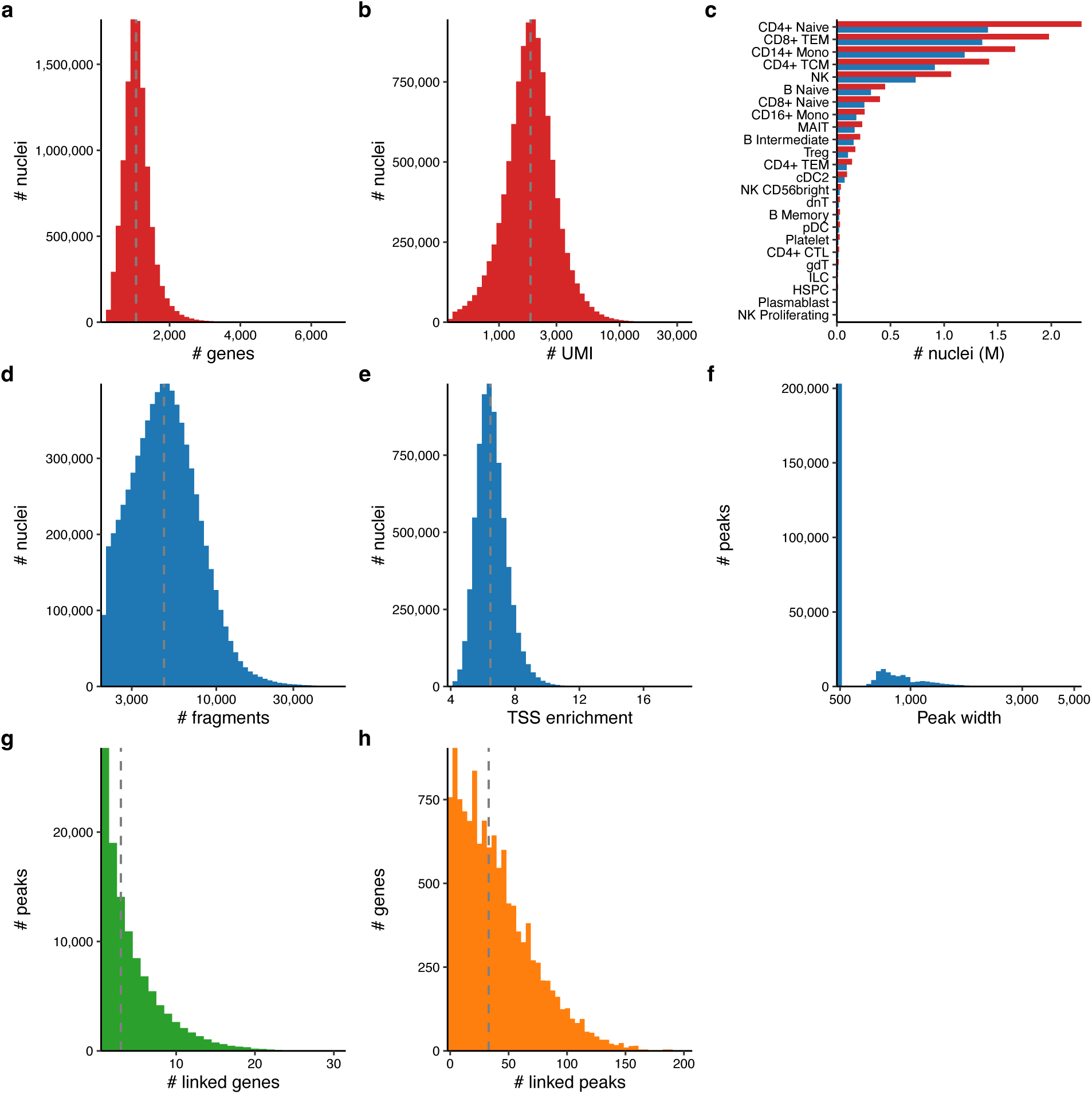
Quality control metrics and cell type composition of snRNA-seq and snATAC-seq atlases a,b, The number of genes (**a**) and the number of UMIs (**b**) per nucleus in the snRNA-seq atlas (10,612,211 nuclei from 1,108 donors). **c**, The number of nuclei for each level 2 (L2) cell types in the snRNA-seq (red) and snATAC-seq (blue). **d,e**, The number of fragments (**d**) and TSS enrichment (**e**) per nucleus in the snATAC-seq atlas (7,100,899 high-quality from 1,093 donors). **f**, Distribution of peak widths across 297,924 consensus peaks. L2 cell-type-specific, fixed-width (500bp) peaks called by MACS2 were merged using iterative overlap peak merging (see **Methods**). **g,h**, The number of linked genes per peak (**g**) and the number of linked peaks per gene (**h**). The dotted vertical line represents the median of the distribution.

**Extended Data Fig. 3.**
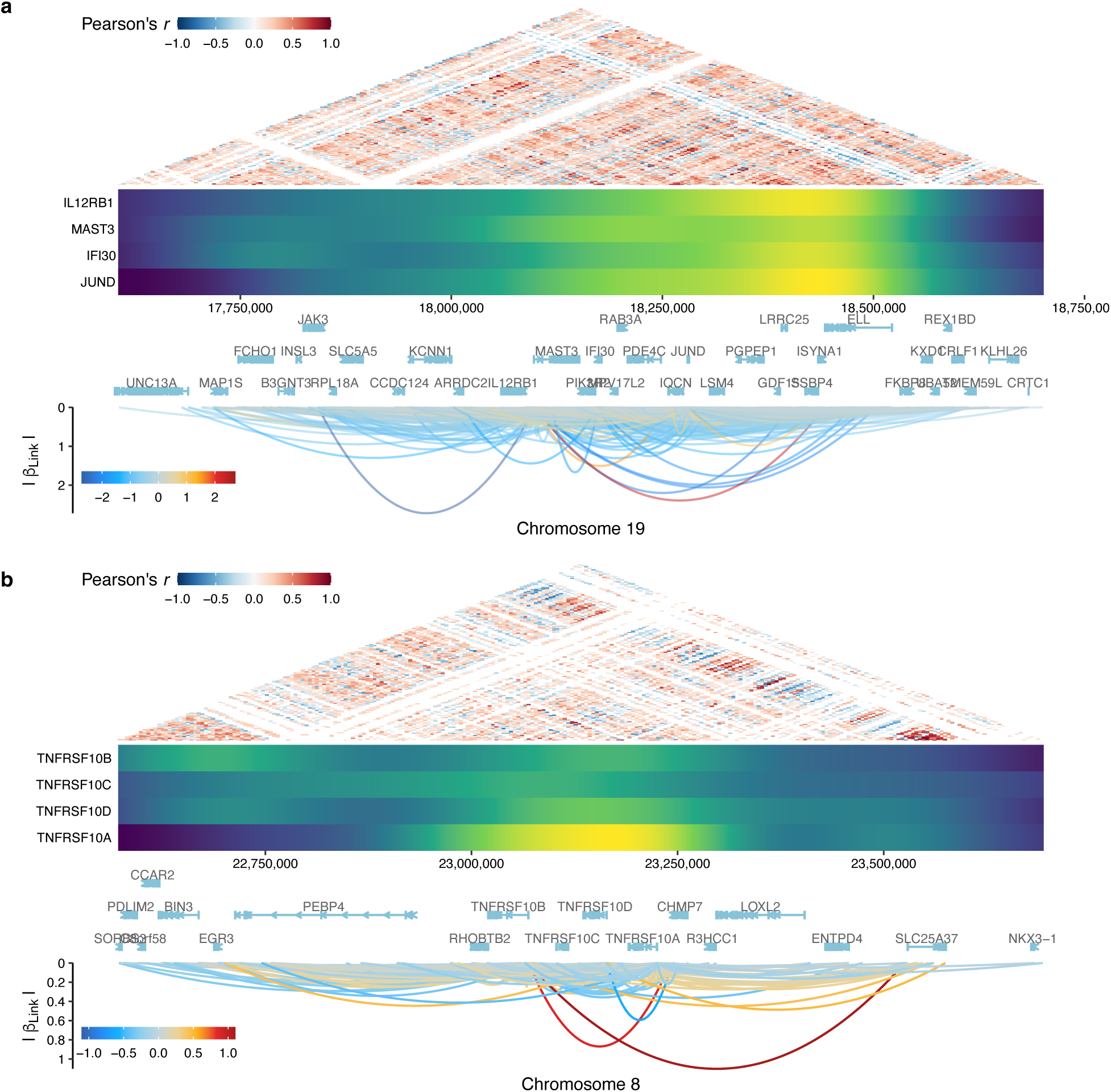
Regulatory hotspots at gene clusters with extensive peak–gene links. Extensive peak–gene links were observed at 19p13.11 containing zinc finger clusters (**a**, *IFI30, JUND, MAST3,* and *IL21RB1*) and 8p21.3 containing TRAIL receptors (**b**, *TNFRSF10A–D*). The top heatmap represents co-accessibility of peaks in this region with significant Pearson correlation (FDR < 0.05, see **Methods**). The middle panel represents the density of peaks linked to each target gene, representing regulatory hotspots of each gene. The bottom panel represents significant peak–gene links with target genes, where color represents effect sizes of links and the y-axis represents the corresponding absolute value.

**Extended Data Fig. 4.**
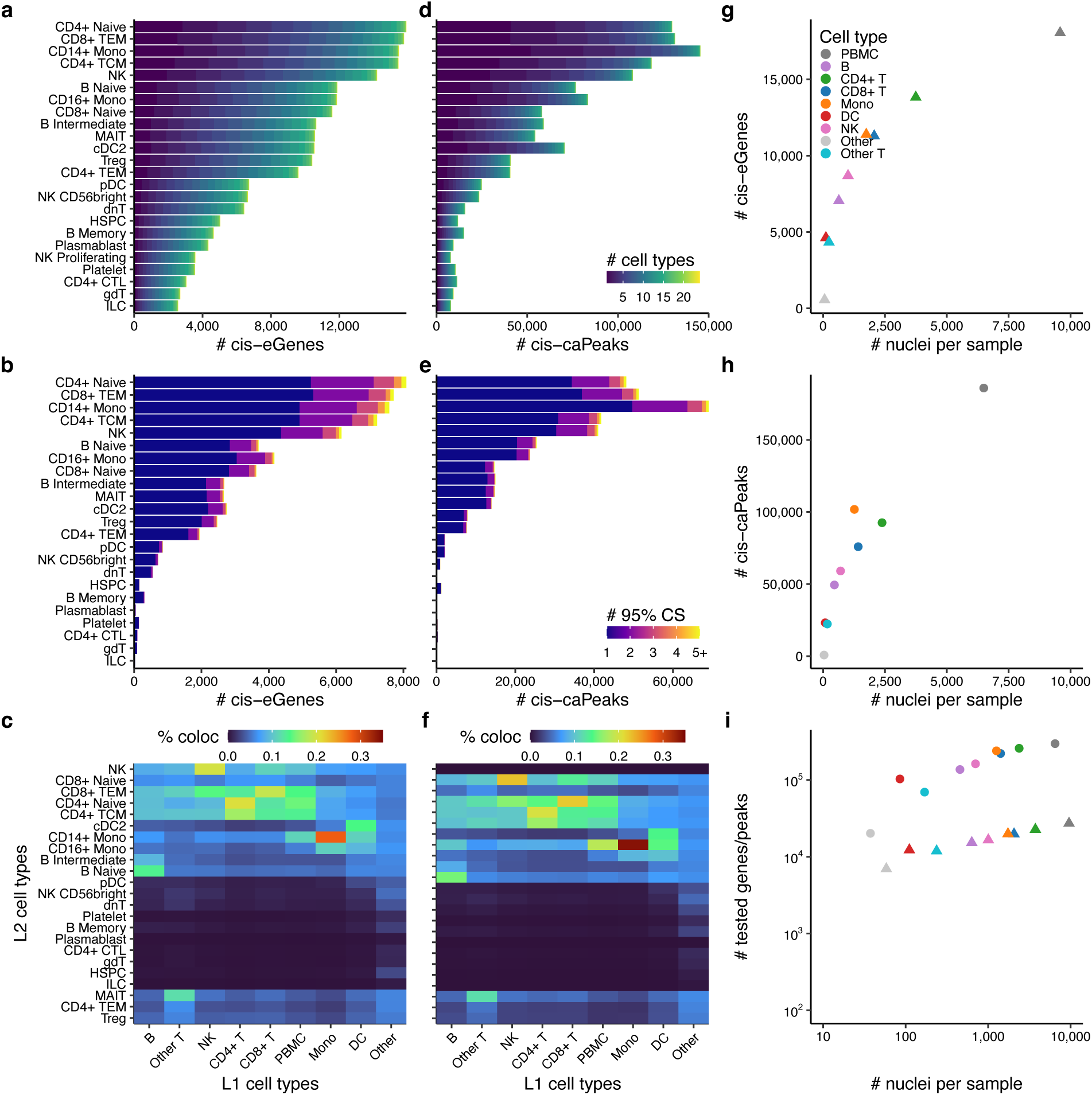
Genetic architecture of molQTLs at fine-grained cell types a,d, The number of significant *cis*-eGenes (**a**) and *cis*-caPeaks (**d**) for each level 2 (L2) cell type (FDR < 0.05; see **Methods**). Color represents the number of cell types that each eGene or caPeak is significantly detected for. **b,e**, The number of *cis*-eGenes (**b**) and *cis*-caPeaks (**d**) with fine-mapped 95% CSs. Color represents the number of 95% CSs for each *cis*-eGene or *cis*-caPeak. **c,f**, Colocalization rates of *cis*-eQTL CSs (**c**) and *cis*-caQTL CSs (**f**) between level 1 (L1) and L2 cell types. Color represents the colocalization rate. The x-and y-axes in both panels were aligned based on the heatmap clustering in eQTLs (**c**) for direct comparison. **g–i**, The relationship between the number of *cis*-eGenes (**g**), caPeaks (**h**), and tested genes or peaks (**i**) and the number of nuclei per sample. Color represents different L1 cell types and shape represents either caQTL or eQTL.

**Extended Data Fig. 5.**
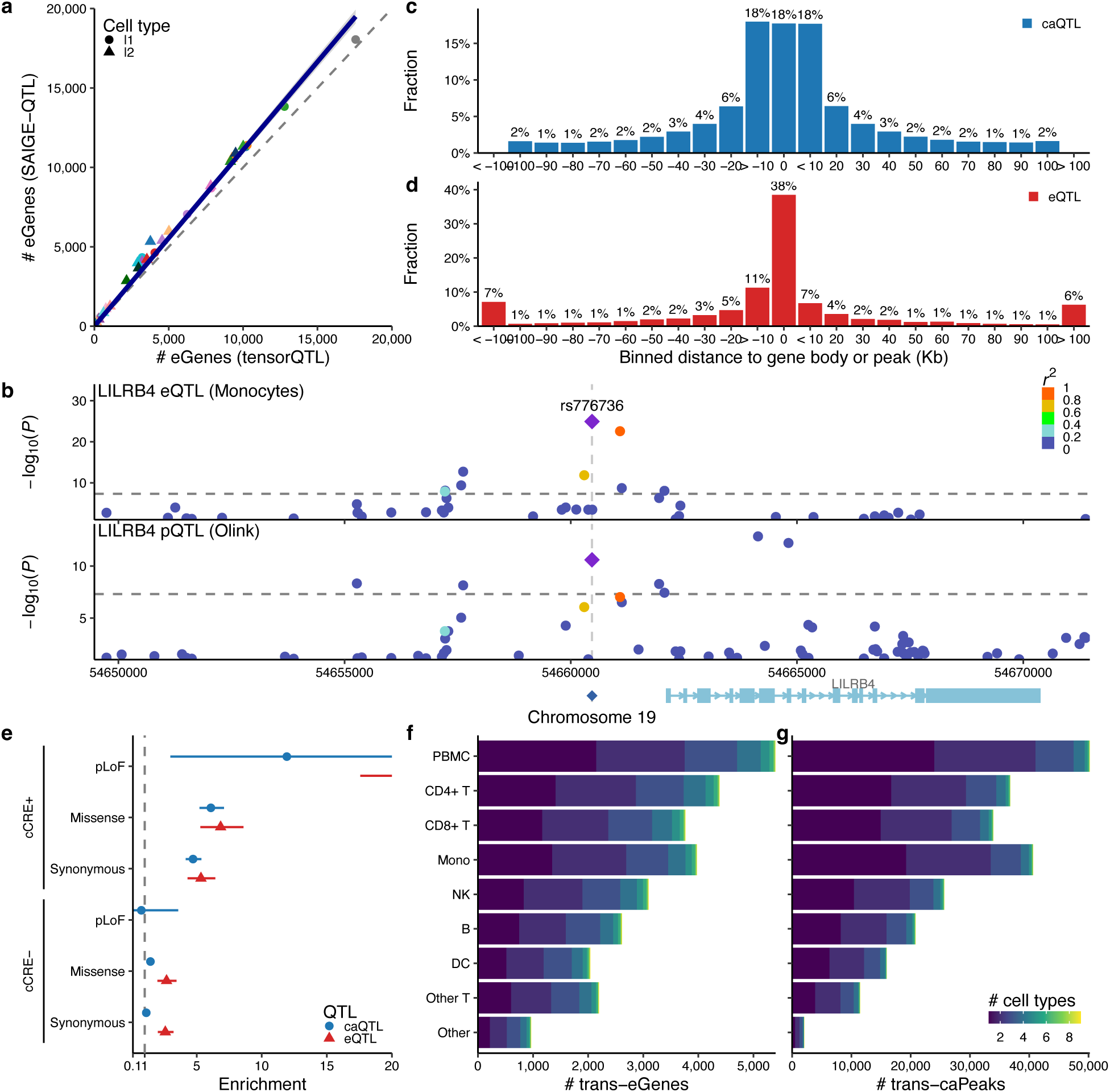
Power comparison between SAIGE-QTL and tensorQTL, functional characterization, and *trans*-QTL analyses. a, Comparison of the number of *cis*-eGenes between SAIGE-QTL and tensorQTL (ACAT FDR < 0.05, see **Methods**). The colored line represents fitted slopes, whereas the dotted slope represents Y = X. **b**, LocusZoom plot of *cis*-eQTL effects for *LILRB4* expression in monocytes and *cis*-pQTL effects for LILRB4 protein levels in plasma. A lead variant, rs776736, is shown as a diamond. Color represents *r*^2^ to the lead variant. Horizontal dotted line represents the genome-wide significance threshold (*P* < 5.0 × 10^-8^). **c,d,** The distribution of fine-mapped eQTL (**c**) or caQTL (**d**) variants around transcription start site (TSS) or peak summit, respectively. Instead of density shown in Fig. 2g, distance to TSS or peak summit is binned by 10 Kbp. **e**, Functional enrichment of high-PIP (> 0.9) caQTL or eQTL coding variants, stratified by cCRE status. Enrichment is defined as a risk ratio of being fine-mapped and being in an annotation (see **Methods**). Error bars represent 95% confidence intervals. **f,g**, The number of significant *trans*-eGenes (**a**) and *trans*-caPeaks (**c**) for each L2 cell type (*P* < 5.0 × 10^-8^; see **Methods**). Color represents the number of cell types that each eGene or caPeak is significantly detected for.

**Extended Data Fig. 6.**
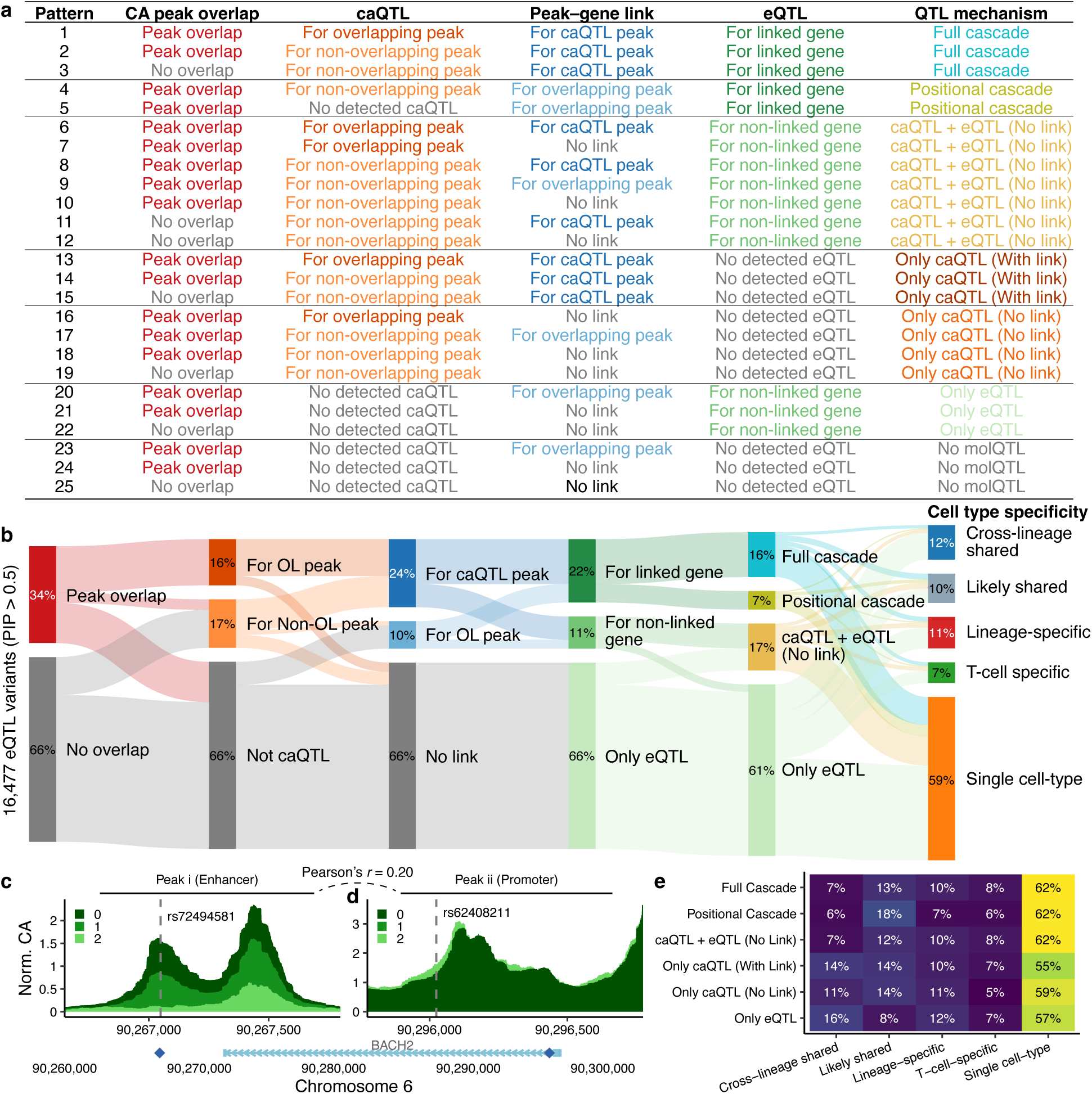
CASCADE classification framework a, Classification scheme showing 25 mutually exclusive patterns based on i) chromatin accessibility (CA) peak overlap,ii) caQTL, iii) peak–gene links, and iv) eQTL, grouped into six mechanistic categories (see **Methods**). **b**. Sankey diagram of the QTL mechanism classifications of 16,477 fine-mapped eQTL variants (PIP > 0.5). For each layer, nodes represent proportions of the fine-mapped molQTL variants. Edges between nodes in each layer represent the fraction of variants shared between nodes. **c,d**, Full regulatory cascade example at *BACH2*, showing rs72928038 regulating chromatin accessibility at peak i (**c**, enhancer), which exhibits significant co-accessibility with peak ii (**d**, promoter). Sequence coverage (CA) is normalized per one million fragments. Color represents the number of alternative alleles of rs72928038, which is near complete linkage disequilibrium with rs62408211 (*r*^2^ = 0.99). Points shown at the bottom as a diamond represent the locations of rs72928038 and rs62408211 at the *BACH2* locus, respectively. **e**, Cell type specificity distribution across the QTL mechanism categories. Color and text represent the fraction for each category.

**Extended Data Fig. 7.**
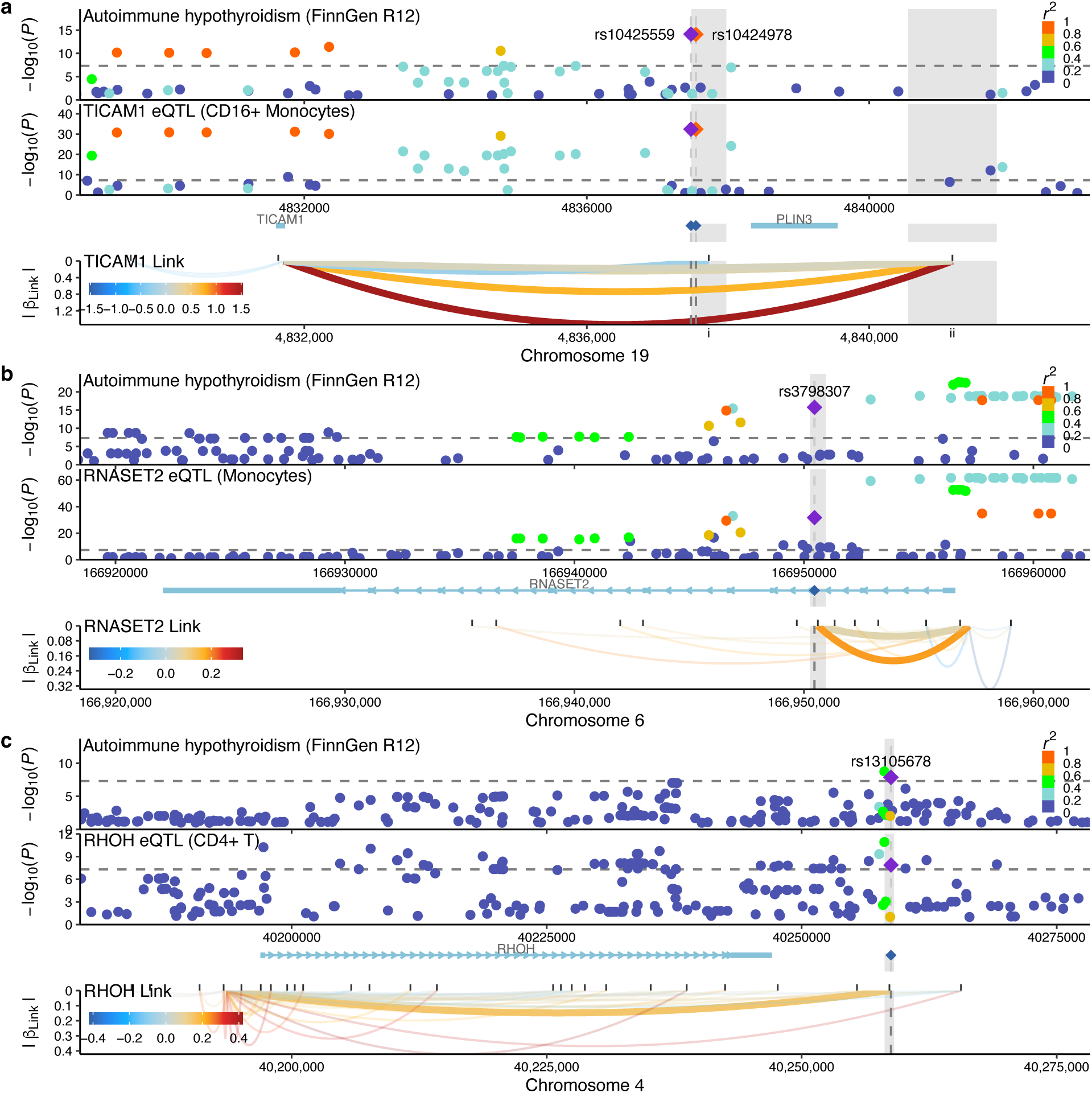
Full cascade regulatory variants for autoimmune hypothyroidism. LocusZoom plots of full cascade variants at *TICAM1* (**a**), *RNASET2* (**b**), and *RHOH* (**c**) loci, colocalizing with autoimmune hypothyroidism (AIHT). For each locus, the top panel represents a LocusZoom plot of AIHT in FinnGen. The middle panel represents *cis*-eQTL effects of individual genes in our dataset. The bottom panel represents peak–gene links between chromatin accessibility peaks and gene expression across cell types. Each curved line represents a link, where the bold line represents peaks involved in the cascade mechanisms. Color represents effect sizes of peak–gene links, whereas Y-axis represents the corresponding absolute value. Marginal rugs at the top represent CA peak locations. Linked peak locations are highlighted as a grey area across panels.

**Extended Data Fig. 8.**
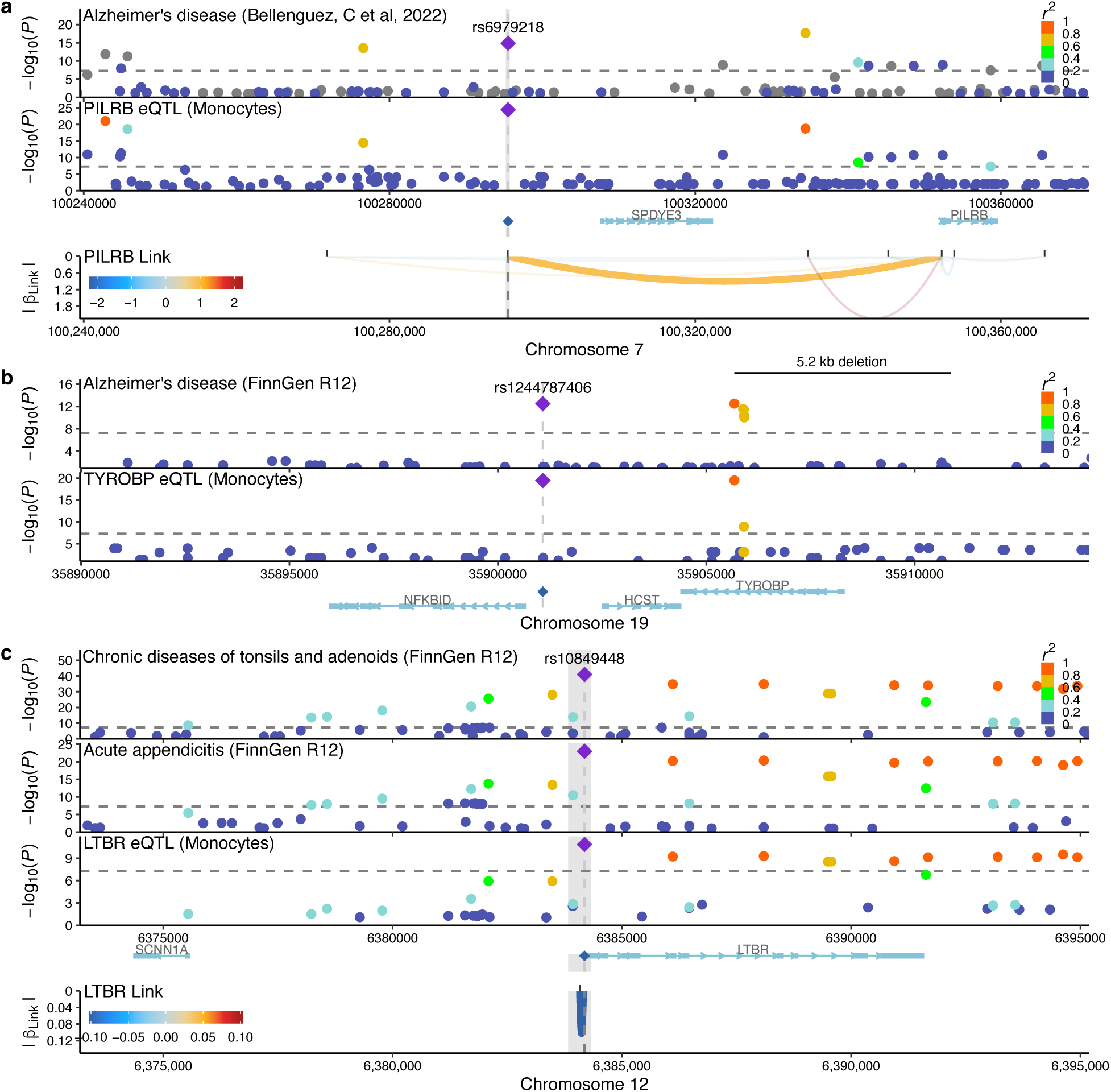
Regulatory cascade mechanisms underlying diseases in non-blood tissues. LocusZoom plots of *PILRB* (**a**), TYROBP (**b**), and LTBR (**c**) loci, colocalizing with complex diseases. For each locus, the top panel represents a LocusZoom plot of AIHT in FinnGen. The middle panel represents *cis*-eQTL effects of individual genes in our dataset. The bottom panel (except for **b**) represents peak–gene links between chromatin accessibility peaks and gene expression across cell types. Each curved line represents a link, where the bold line represents peaks involved in the cascade mechanisms. Color represents effect sizes of peak–gene links, whereas Y-axis represents the corresponding absolute value. Marginal rugs at the top represent CA peak locations. Linked peak locations are highlighted as a grey area across panels.

**Extended Data Fig. 9.**
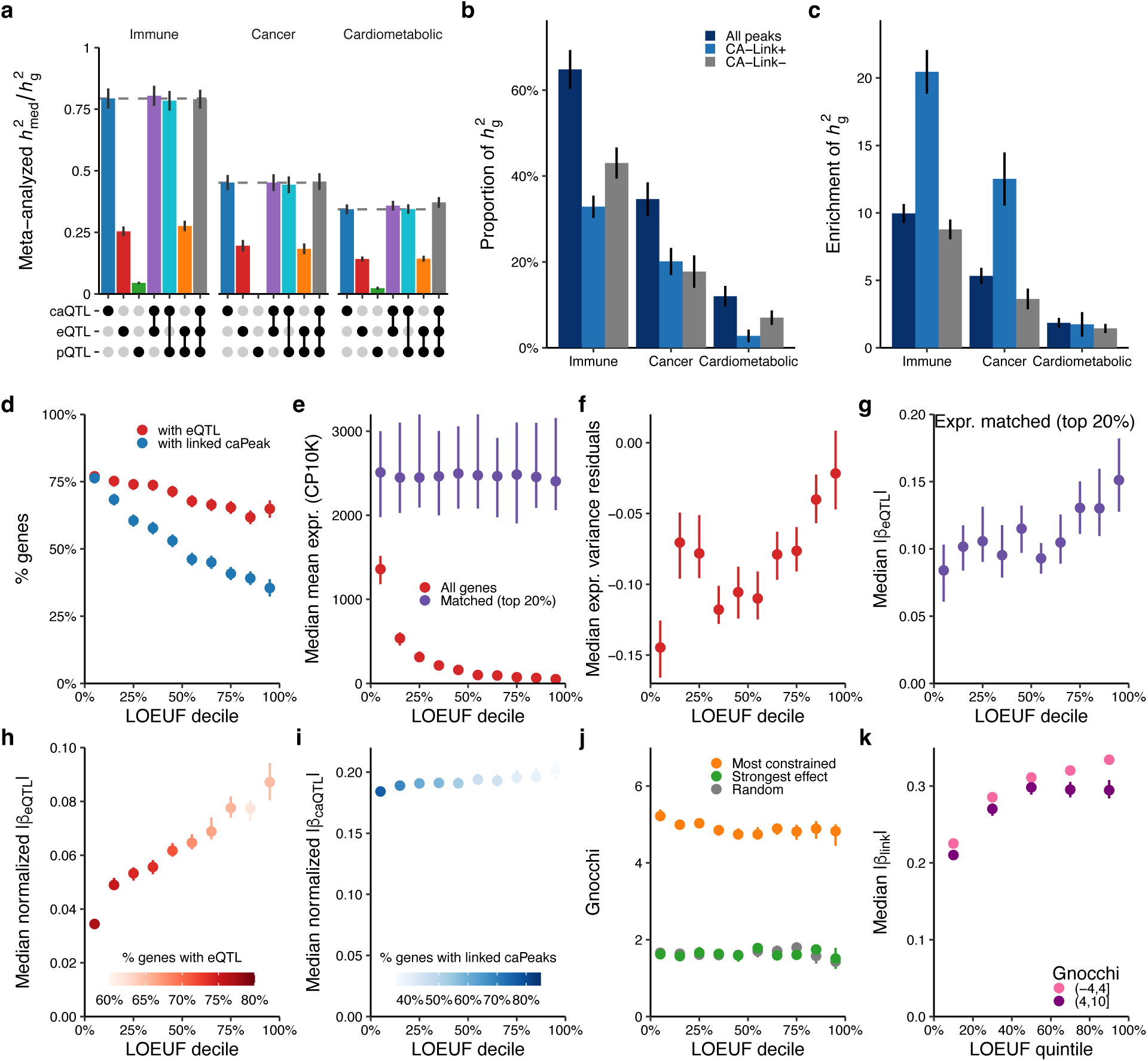
QTL-mediated disease heritability, and multi-layered regulatory buffering a, Estimated proportion of disease heritability mediated by molecular traits. For immune, cancer, and cardiometabolic traits, MESC-estimated *h*^2^med / *h*^2^g within each trait category are meta-analyzed using random-effect model (**Methods**). The x-axis represents the combination of molQTLs for each estimate. The error bar represents standard errors. Horizontal dotted lines represent the point estimate for caQTL in each trait category. **b,c**, Estimated proportion of disease heritability (b) and heritability enrichment (c), stratified by chromatin accessibility (CA) peak with links (CA–Link+) and without (CA–Link–). **d**, The relationship between the fraction of genes with eQTL or linked peaks affected by caQTL and loss-of-function observed/expected upper bound fraction (LOEUF) deciles. **e**, The median of normalized mean expression (counts per 10,000 nuclei, CP10K) across LOEUF deciles, computed for all genes and expression-matched highly expressed (top 20%) genes across the deciles. **f**, The median of expression variance residuals, corrected by mean expression, across LOEUF deciles (see **Methods**). **g**, The median absolute *cis*-eQTL effect sizes for expression-matched genes across LOUEF deciles. **h,i**, The median MAF-normalized absolute effect sizes for *cis*-eQTLs (**h**) and *cis*-caQTLs (**i**). Color represents the fraction of genes with either eQTL (blue) or linked peaks affected by caQTL (red) for each decile. **j**, The distribution of median genomic non-coding constraint of haploinsufficient variation (Gnocchi), computed for most constrained, strongest effect, or randomly selected peaks. **k**, The median absolute link effect sizes, stratified by both peak and gene constraints. LOEUF quintiles, not deciles, were used to account for a limited number of genes (see **Methods**). Gnocchi threshold of 4 was used to group constrained and unconstrained peaks. Error bars represent 95% confidence intervals.

**Extended Data Fig. 10.**
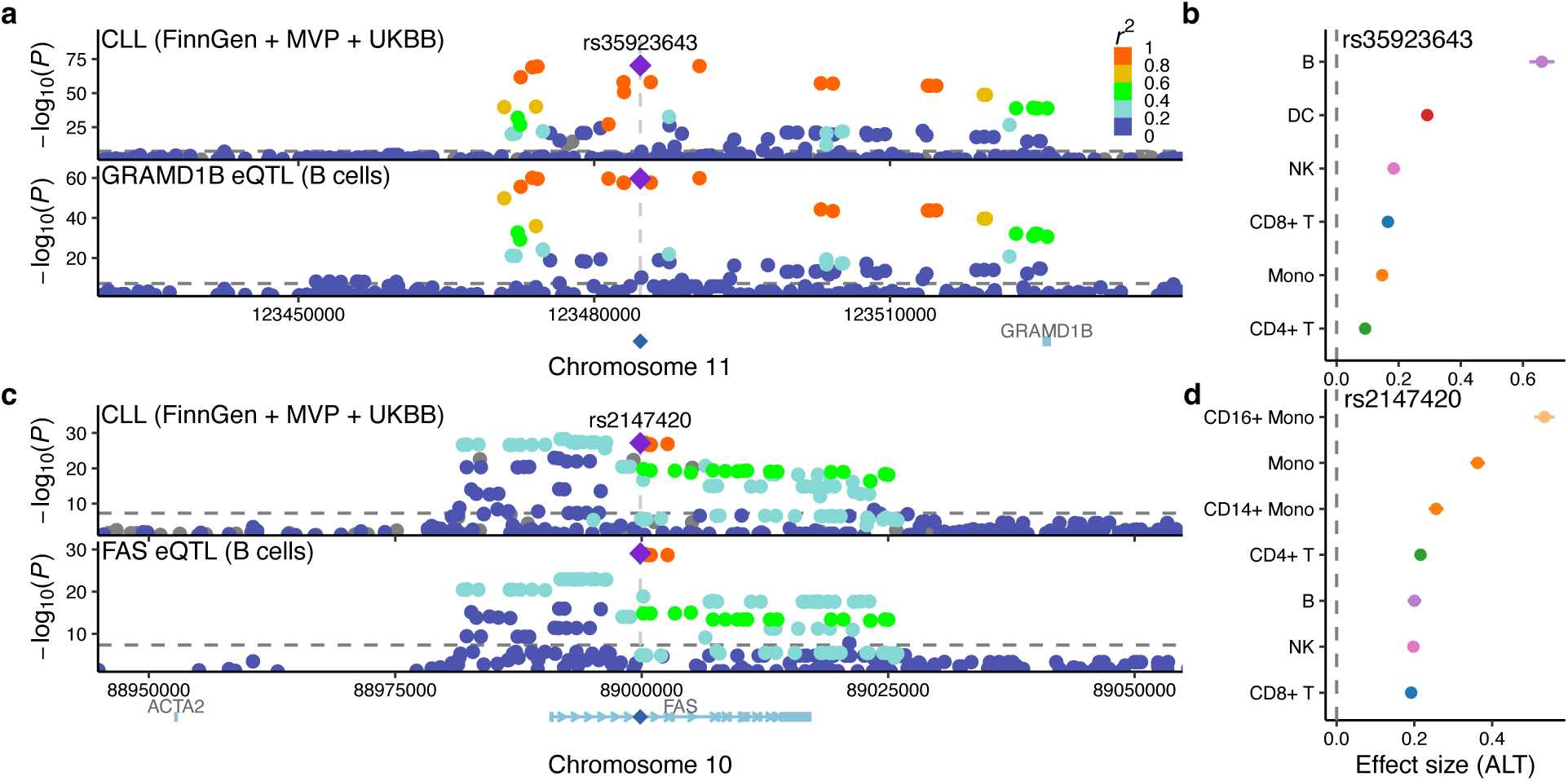
**Constrained genes with substantial *cis*-eQTL effects a,c**, LocusZoom plots of *GRAMD1B* (**a**) and *FAS* (**c**) loci, colocalizing with chronic lymphocytic leukemia (CLL). For each locus, the top panel represents a LocusZoom plot of CLL in the FinnGen + MVP + UKBB meta-analysis. The bottom panel represents *cis*-eQTL effects of *GRAMD1B* or *FAS* expression in B cells. A lead variant for each locus is shown as a diamond. Color represents *r*^2^ to the lead variant. Horizontal dotted line represents the genome-wide significance threshold (*P* < 5.0 × 10^-8^). **b,d**, Forest plots of rs35923643 (**b**) and rs2147420 (**d**) for *GRAMD1B* or *FAS* expression, respectively, across cell types.

